# The Effects of Serotonergic systems on Cognitive Flexibility and Perseverative Thinking: a comparison between SSRI, classical psychedelics, and acute tryptophan depletion in a Multilevel Meta-Analysis

**DOI:** 10.64898/2026.06.18.26355974

**Authors:** Roi Basch, Maya Cohen, Leehe Peled-Avron

## Abstract

**Background:** Serotonin has been implicated in cognitive flexibility and pathological perseverative thinking (PT), including rumination, worry, and obsessions. However, evidence remains fragmented across pharmacological manipulations, clinical populations, and outcome measures. This multilevel meta-analysis examined whether serotonergic interventions influence PT and cognitive flexibility.

**Methods:** Preregistered and following PRISMA guidelines, we synthesized studies investigating three classes of serotonergic manipulations: acute tryptophan depletion (ATD), serotonin elevation via selective serotonin reuptake inhibitors (SSRIs), and classic serotonergic psychedelics. Three multilevel random-effects meta-analyses with cluster-robust variance estimation were conducted: (A) effects of ATD on cognitive flexibility (N = 266; 10 effect sizes), (B) effects of serotonin elevation on cognitive flexibility (N = 654; 15 effect sizes), and (C) effects of serotonin elevation on pathological perseverative thinking (N = 1,100; 20 effect sizes). Across analyses, the total sample comprised 2,030 participants and 45 effect sizes.

**Results:** ATD did not significantly impair cognitive flexibility (g = 0.15, 95% CI [-0.07, 0.38], p =.23), and no moderation by task type, sex, or age was observed. Serotonin elevation similarly did not improve cognitive flexibility (g =-0.07, 95% CI [-0.36, 0.22], p =.63), with no significant performance differences emerging between SSRIs, classical psychedelics, or tryptophan enrichment. In contrast, serotonin elevation was associated with a significant medium-to-large reduction in perseverative thinking (g =-0.58, 95% CI [-0.76,-0.41], p <.001). Notably, while both pharmacological classes effectively reduced cognitive rigidity, SSRIs demonstrated a marginally smaller magnitude of symptom reduction compared to acute psilocybin interventions (p =.081). Furthermore, samples with a higher proportion of female participants showed larger reductions in perseverative thinking (β =-1.86, p =.014), while worries exhibited marginally smaller reductions relative to obsessions (β = 0.42, p =.055). Publication bias tests were non-significant across analyses.

**Conclusions:** Serotonergic interventions robustly reduce perseverative thinking but do not consistently alter performance on laboratory measures of cognitive flexibility. These findings suggest that serotonin may influence cognitive-emotional rigidity and the subjective experience of repetitive thought more strongly than objective executive task performance. The dissociation between task-based and phenomenological outcomes aligns with contemporary models of serotonergic plasticity and highlights perseverative thinking as a potentially transdiagnostic therapeutic target of serotonergic interventions.

---

Perseverative thinking (PT), often operationalized in the literature as perseverative cognition, refers to the repeated or chronic cognitive representation of stressors that persists well beyond the presence of an immediate threat (Brosschot et al., 2006). These repetitive, difficult-to-control patterns of thought lose their adaptive value and become maladaptive when they prolong emotional and physiological activation (Brosschot et al., 2006). As a broad construct, PT encompasses several closely related pathological patterns (PPTP), including worry, rumination and obsessions (Brosschot et al., 2006; Hallion et al., 2022). Although these forms differ in content and temporal orientation, they share several core characteristics: repetitiveness, reduced controllability, attentional fixation, and impaired ability to disengage from internal thoughts (Ehring & Watkins, 2008).

Phenomenologically, rumination is typically past-oriented and focused on negative self-referential processing (Nolen-Hoeksema et al., 2008), worry tends to be future-oriented and associated with uncertainty and threat anticipation (Borkovec & Inz, 1990), and obsessions are intrusive, often ego-dystonic thoughts commonly observed in obsessive-compulsive disorder (Purdon & Clark, 1999). However, cognitive testing reveals that these distinct phenotypic expressions are driven by an identical underlying deficit: an impaired capacity for cognitive control, specifically characterized by a reduced ability to shift attention away from internally generated negative representations and a marked decline in mental flexibility (Hsu et al., 2015; Miyake & Friedman, 2012; Watkins, 2008).

PPTP represent a core transdiagnostic vulnerability factor that drives the development, severity, and maintenance of a vast spectrum of psychopathology, including major depressive disorder, generalized anxiety disorder, OCD, post-traumatic stress disorder, and eating disorders (Ehring & Watkins, 2008; Harvey et al., 2004). Despite its profound clinical significance across these conditions, PT has proven exceptionally resilient to standard psychiatric interventions. Traditional pharmacological and psychotherapeutic protocols frequently alleviate acute affective symptoms without fully dismantling the rigid, underlying repetitive cognitive architecture itself, resulting in high rates of clinical relapse (Stenzel et al., 2025; Watkins, 2008). This persistent therapeutic barrier suggests that current treatments may only be targeting the downstream symptomatic surface of the problem. Instead, the fundamental mechanism driving this treatment resistance may stem from deeper, foundational failures in basic cognitive control - specifically, distinct impairments in cognitive flexibility and working memory updating that keep the mind structurally incapable of disengaging from repetitive internal loops (Gabrys et al., 2018; Miyake & Friedman, 2012).

Robust correlational and meta-analytic data consistently establish a strong negative relationship between PPTP and cognitive flexibility. Cognitive flexibility (CF) is broadly defined as the executive capacity to adaptively switch thoughts, perspectives, or behavioral strategies in response to changing environmental contingencies or shifting internal demands (Diamond, 2013; Miyake & Friedman, 2012).; Specifically, significantly diminished CF consistently predicts a higher frequency, duration, and severity of transdiagnostic PPTP, across both clinical and non-clinical populations (Davis & Nolen-Hoeksema, 2019; Snyder et al., 2015; Kertz et al., 2016; Lee & Orsillo, 2014).

Underneath the hood, this strong behavioral connection between CF and PPTP is driven by highly specific psychological and neurobiological mechanisms. Psychologically, the transition from healthy processing to rigid PT reflects a breakdown in the core sub-components of executive function: attentional shifting and working memory updating (Miyake & Friedman, 2012). When a person exhibits low cognitive flexibility, their working memory may become “captured” by negative, self-relevant information. A potential lack of top-down cognitive control may inhibit updating of working memory or shift attention away from these internal representations and thus, the loop remains active, cementing the perseverative pattern (Joormann & Gotlib, 2010).

Neurobiologically, this cognitive failure is instantiated by an aberrant, poorly coordinated interplay between two large-scale neural networks: the Default Mode Network (DMN), which governs internally focused, self-referential cognition, and the Frontoparietal Control Network (FCN), which acts as the primary hub for top-down executive oversight (Kaiser et al., 2015; Menon, 2011). In cognitively flexible individuals, the FCN exerts rapid, adaptive top-down control over the DMN, effectively down-regulating or disengaging from self-referential circuits when internal representations conflict with current task goals (Andrews-Hanna et al., 2014). In individuals prone to pathological PT, however, this dynamic coordination breaks down. Rather than executing flexible network transitions, the brain exhibits an altered functional coupling marked by a failure of the FCN to exert sufficient top-down inhibition over hyper-reactive, self-relevant DMN hubs (Hamilton et al., 2011; van Oort et al., 2022). This structural and functional network dysregulation operates as a core transdiagnostic vulnerability; multivariate neural representations of future-focused worry and past-focused rumination show massive overlapping footprints across these exact frontoparietal and default-mode circuits, pinning network inflexibility as the common engine driving pathological perseveration across the psychopathological continuum (Doucet et al., 2020; Puccetti et al., 2025).

The dynamic, real-time coordination of these large-scale network interactions is fundamentally governed by ascending neuromodulatory systems that tune cortical excitability and gate regional information flow. Foremost among these is serotonin (5-hydroxytryptamine; 5-HT), a monoaminergic neurotransmitter synthesized within the brainstem raphe nuclei that projects extensively to both frontoparietal and default mode network hubs (Hahn et al., 2012). Serotonergic signaling modulates target cortical circuits through a dense architecture of distinct receptor subtypes, with the G-protein-coupled 5-HT_1A_and 5-HT_2A_ receptors highly enriched on the apical dendrites of pyramidal neurons in the prefrontal neocortex (Albert et al., 2014; Celada et al., 2004). Postsynaptic 5-HT_1A_ heteroreceptors typically mediate local inhibition, whereas 5-HT_2A_ activation functions as the primary excitatory driver of frontocortical networks, critically modulating cognitive control, working memory, and attentional allocation (Albert et al., 2014). Because of this widespread modulatory footprint, systematic dysregulation of the serotonergic system has has been associated with major depression, anxiety disorders, and obsessive-compulsive conditions - pathologies commonly characterized by cognitively rigid, repetitive behavior (Cools et al., 2011)

Emerging computational and neurobiological models suggest that serotonin contributes to these diverse psychopathologies not by merely regulating raw affective tone, but by acting as a fundamental signal for environmental uncertainty, plasticity, and behavioral updating (Carhart-Harris & Nutt, 2017; Cools et al., 2011). Under normal conditions, healthy serotonin transmission allows an organism to dynamically adapt to changing environmental contingencies, with midbrain serotonin neurons tracking prediction errors to signal when deeply ingrained internal representations or schemas must be revised (Matias et al., 2017). When serotonergic functioning is blunted or compromised, this capacity for active cognitive adaptation breaks down. Lacking the necessary neuromodulatory signaling to drive synaptic plasticity and disengage from non-task-relevant internal representations, the brain defaults to automated, rigid cognitive styles. This neurochemical failure may manifest as the unyielding, repetitive cognitive loops characteristic of pathological PT, whereas agonists to serotonin 2A receptors (psilocybin) were found to restore the neuroplastic capacity associated with adaptive shifting behavior (Carhart-Harris & Nutt, 2017).

This theoretical framework raises the possibility that serotonergic interventions may influence pathological perseverative thinking patterns by altering cognitive flexibility. If serotonin enhances the capacity to update beliefs, shift attention, and adapt behaviour, serotonergic manipulations may reduce perseverative cognition indirectly through improvements in flexibility-related processes. However, evidence regarding this relationship remains fragmented across different pharmacological manipulations and experimental paradigms.

To systematically quantify how bidirectional alterations in serotonergic tone relate to executive control and PT, this meta-analysis evaluates studies utilizing three distinct classes of pharmacological and dietary serotonin manipulations: acute tryptophan depletion (ATD), selective serotonin reuptake inhibitors (SSRIs), and classic serotonergic psychedelics. ATD serves as an experimental model for transiently lowering central serotonin levels; by administering a concentrated amino acid mixture lacking tryptophan, this paradigm forces hepatic protein synthesis and activates transport competition at the blood-brain barrier, rapidly restricting precursor availability and central 5-HT synthesis within the brainstem raphe nuclei (Mendelsohn et al., 2009; van Donkelaar et al., 2011). Conversely, SSRIs represent the primary clinical mechanism for augmenting endogenous serotonergic transmission; these compounds acutely elevate extracellular 5-HT concentration throughout frontocortical and subcortical interstitial spaces by competitively inhibiting the presynaptic serotonin transporter (SERT) and preventing neurotransmitter reuptake (Hiemke & Härtter, 2000). Finally, classic serotonergic psychedelics (e.g., psilocybin, LSD) introduce a direct exogenous mechanism, altering neural signaling primarily by acting as high-affinity agonists or partial agonists at the cortical 5-HT_2A_ receptor (Nichols, 2016). Unlike the endogenous upregulation seen with SSRIs, direct 5-HT_2A_ receptor activation by psychedelics uniquely alters deep layer V pyramidal neuron excitability, disrupting rhythmic macro-scale network synchronization and temporarily increasing global neural entropy (Carhart-Harris et al., 2014). By grouping the literature into these three physiological archetypes, this meta-analysis establishes a comparative framework to examine how distinct modes of serotonergic manipulation differentially impact PT and cognitive flexibility.

## The Current Study

There are several gaps in the literature: (1) Despite accumulating evidence on serotonin’s role in PT and cognitive flexibility, the existing empirical literature remains heavily fragmented across disconnected clinical populations, varying pharmacological models, and highly diverse testing paradigms. (2) Direct quantitative comparisons across distinct modes of serotonergic manipulation are currently lacking in the literature, and it is unclear whether disparate pharmacological inputs converge onto identical or distinct aspects of PT. By synthesizing evidence across experimental depletion paradigms, chronic clinical interventions, and acute psychedelic states, this meta-analysis addresses these fragmentation gaps. Rather than testing the already well-established baseline relationship connecting cognitive rigidity to clinical distress, this work evaluates whether distinct classes of serotonergic interventions simultaneously modulate both the objective cognitive structures and the subjective clinical expressions of psychological inflexibility.

To achieve this integration, the present meta-analysis executes three distinct, complementary statistical analyses designed to systematically map how diverse serotonergic manipulations affect these twin dimensions of rigidity (PT and cognitive flexibility).

**Analysis A** addresses the cognitive impact of acute serotonergic reduction: The research question is how does a transient decrease in central serotonin via acute tryptophan depletion affect objective performance on cognitive flexibility measures? We hypothesize a primary main effect wherein ATD significantly impairs objective flexibility outcomes; additionally, we will exploratorily evaluate whether this effect is significantly moderated by biological sex, age, or specific cognitive task type (i.e., feedback-driven reversal learning versus cue-based set-shifting).

**Analysis B** examines the effects of serotonin increase on cognitive flexibility. The research question is how does the elevation of serotonergic activity, driven by either SSRIs or classic psychedelics, affect objective cognitive flexibility measures? Here, we directionalize our primary hypothesis toward an overall enhancement of flexible performance, while treating substance type, exposure timeline parameters (acute vs. chronic schedules), age, and biological sex as exploratory moderators to determine potential boundaries or variations in effect sizes.

Finally, **Analysis C** pivots to the clinical manifestation of PT The research question is how does serotonergic elevation via SSRIs or classical psychedelics alter subjective self-report and clinical expressions of pathological perseverative thinking patterns (PPTP)? We hypothesize that serotonergic enhancement will significantly decrease self-reported and clinical manifestations of PT. Given the clinical heterogeneity across the literature, we will run exploratory moderation analyses across several key clinical and design variables, including the specific phenomenological construct assessed (rumination, worries, vs. obsessions), the choice of pharmacological manipulation (SSRI or psychedelics), biological sex, and age. This comprehensive approach would empirically map which factors systematically alter the magnitude of this hypothesized reduction.

Combined, these three distinct research questions provide a thorough evaluation of the effect of serotonin on PT. Analysis A and B map the chemical boundaries of objective cognitive control, while Analysis C charts the clinical modulation of its symptomatic expression, collectively testing whether optimizing serotonergic functioning serves as a unified pathway for reducing transdiagnostic cognitive and clinical rigidity.

## Methods

This systematic review and meta-analysis was preregistered on the Open Science Framework (OSF; https://osf.io/ugwma) and conducted in accordance with the PRISMA (Preferred Reporting Items for Systematic Reviews and Meta-Analyses) reporting guidelines (Moher et al., 2015).

### Search Strategy and Eligibility Criteria

A comprehensive literature search was performed across PubMed (Medline), PsycINFO, and Scopus databases from their inception through March 2026. The study selection process is visually detailed in the flowchart presented in Figures 1,2. The search utilized the following keyword combinations:

**Figure 1.**
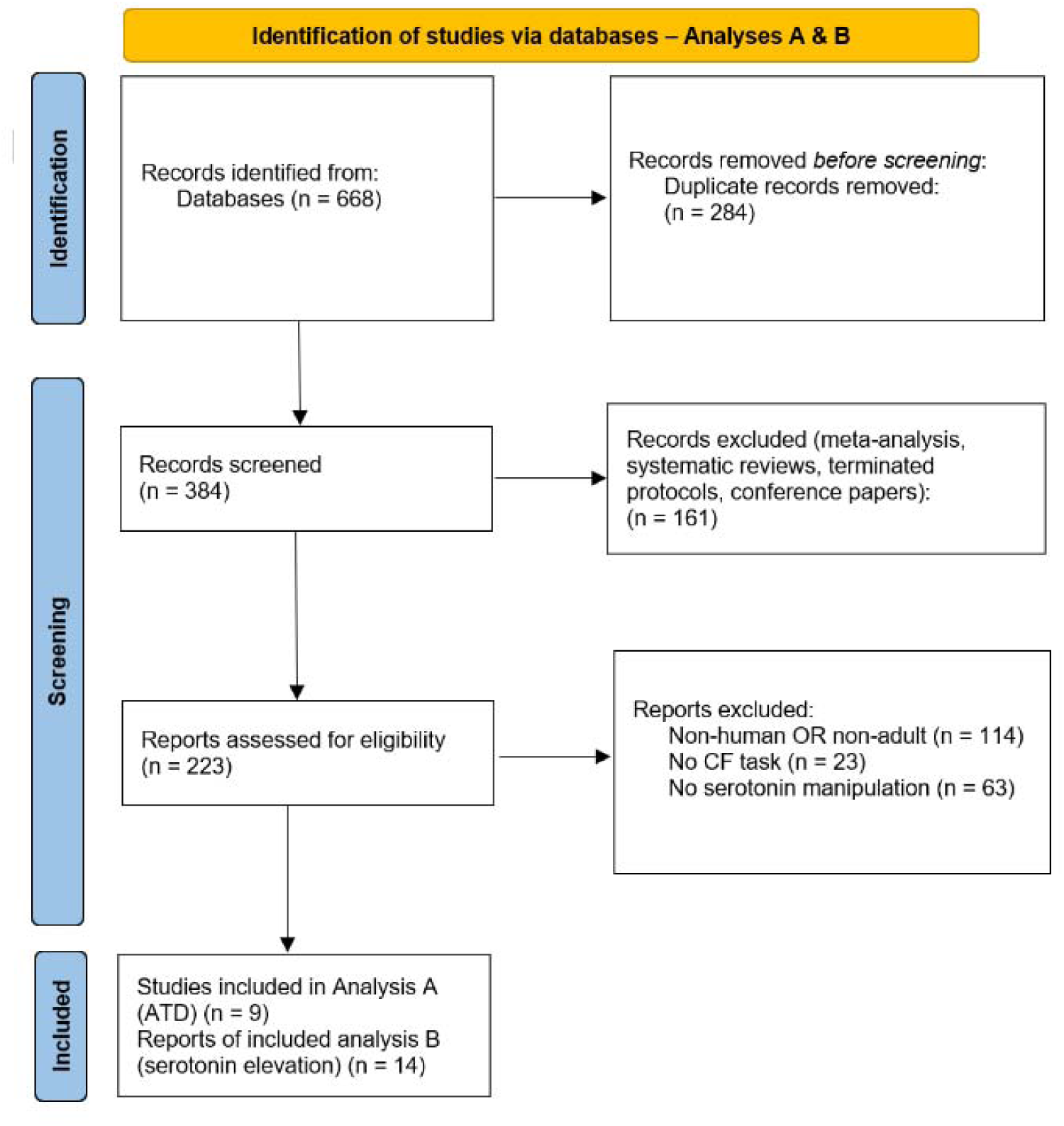
The search and selection procedure for analyses A & B (acute tryptophan depletion and serotonin elevation manipulations effects on cognitive flexibility tasks) to identify studies for inclusion. Template provided by PRISMA (www.prisma-statement.org).

**Figure 2.**
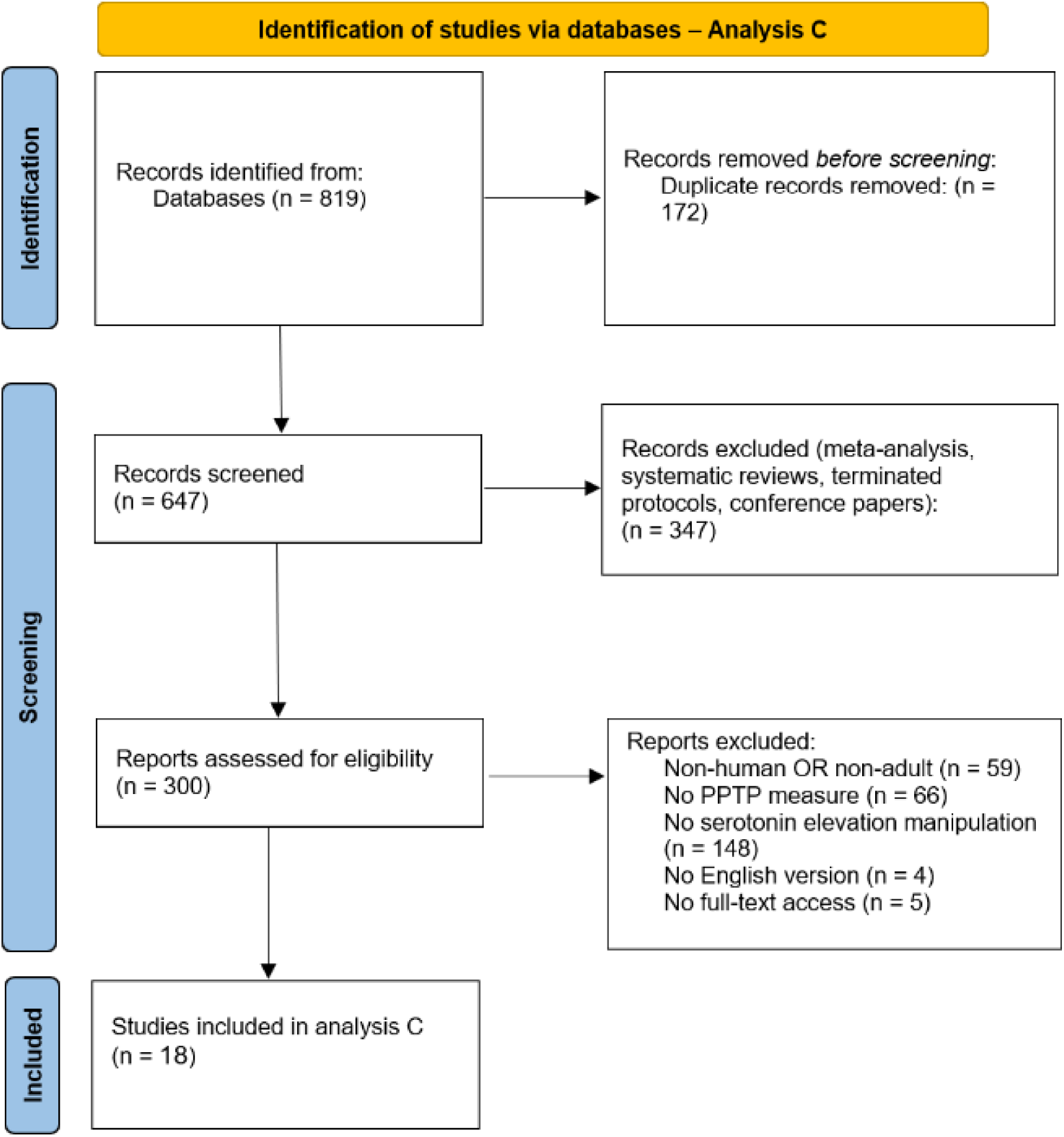
The search and selection procedure for analysis C (Serotonin elevation manipulations effects on PPTP measures) to identify studies for inclusion. Template provided by PRISMA (www.prisma-statement.org).

For analyses A & B:

- “Serotonin” AND “Cognitive flexibility”.

- “SSRI” AND “Cognitive flexibility”

- “Tryptophan depletion” AND “Cognitive flexibility”

- “Psychedelics” AND “Cognitive flexibility”

For analysis C:

- (“Serotonin” OR “SSRI” OR “Tryptophan depletion” OR “Psychedelics”) AND “Rumination”

- (“Serotonin” OR “SSRI” OR “Tryptophan depletion” OR “Psychedelics”) AND “worries”

- (“Serotonin” OR “SSRI” OR “Tryptophan depletion” OR “Psychedelics”) AND “obsession”

The selection and screening of literature for inclusion in this meta-analysis followed a systematic, multi-stage workflow to isolate eligible empirical studies. Following the initial identification of records across the mentioned electronic databases, all duplicate records were algorithmically identified and removed. The remaining unique records were then subjected to a preliminary screening phase based strictly on titles and abstracts to remove non-primary empirical categories, such as reviews, meta-analyses, terminated protocols, comments, letters, editorials, conference proceedings or clearly non-relevant records.

The remaining articles underwent full-text evaluation against explicit inclusion criteria. To meet eligibility, studies were required to be original, peer-reviewed, full-text research articles published in English, utilizing an adult human sample. Critically, the experimental design was required to include both a serotonin manipulation (with no a priori restrictions placed on participant age, sex, compound dosage, frequency, or route of administration) and at least one objective measure of a cognitive flexibility task and/or a validated measure of a perseverative thinking pattern (specifically restricted to rumination, worry, or obsession).

However, to ensure the neurochemical specificity of the synthesized data, eligible manipulations were strictly restricted to interventions targeting the serotonin system in isolation (e.g., ATD, SSRIs, or classic psychedelics); pharmacology utilizing broad, multi-neurotransmitter mechanisms - such as MDMA, atypical antipsychotics, or mixed serotonin-norepinephrine reuptake inhibitors (SNRIs) - were explicitly excluded. Additionally, the reference lists of eligible articles were manually screened to identify any further relevant studies. The demographic characteristics and experimental parameters of the included studies are summarized in Tables 1, 2, and 3.

**Table 1.**
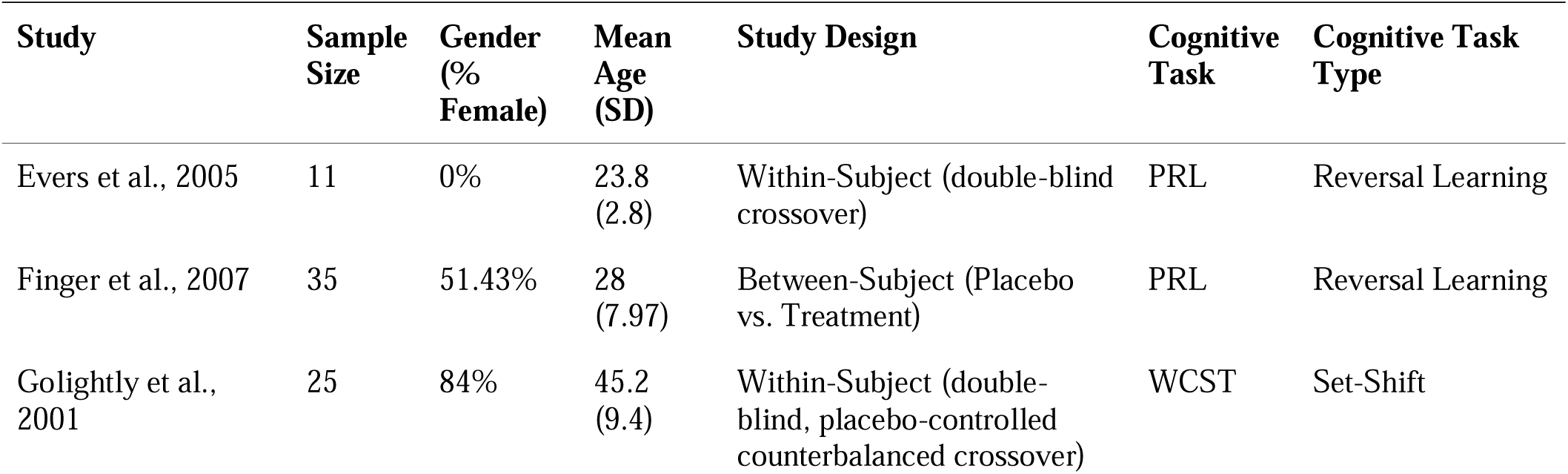
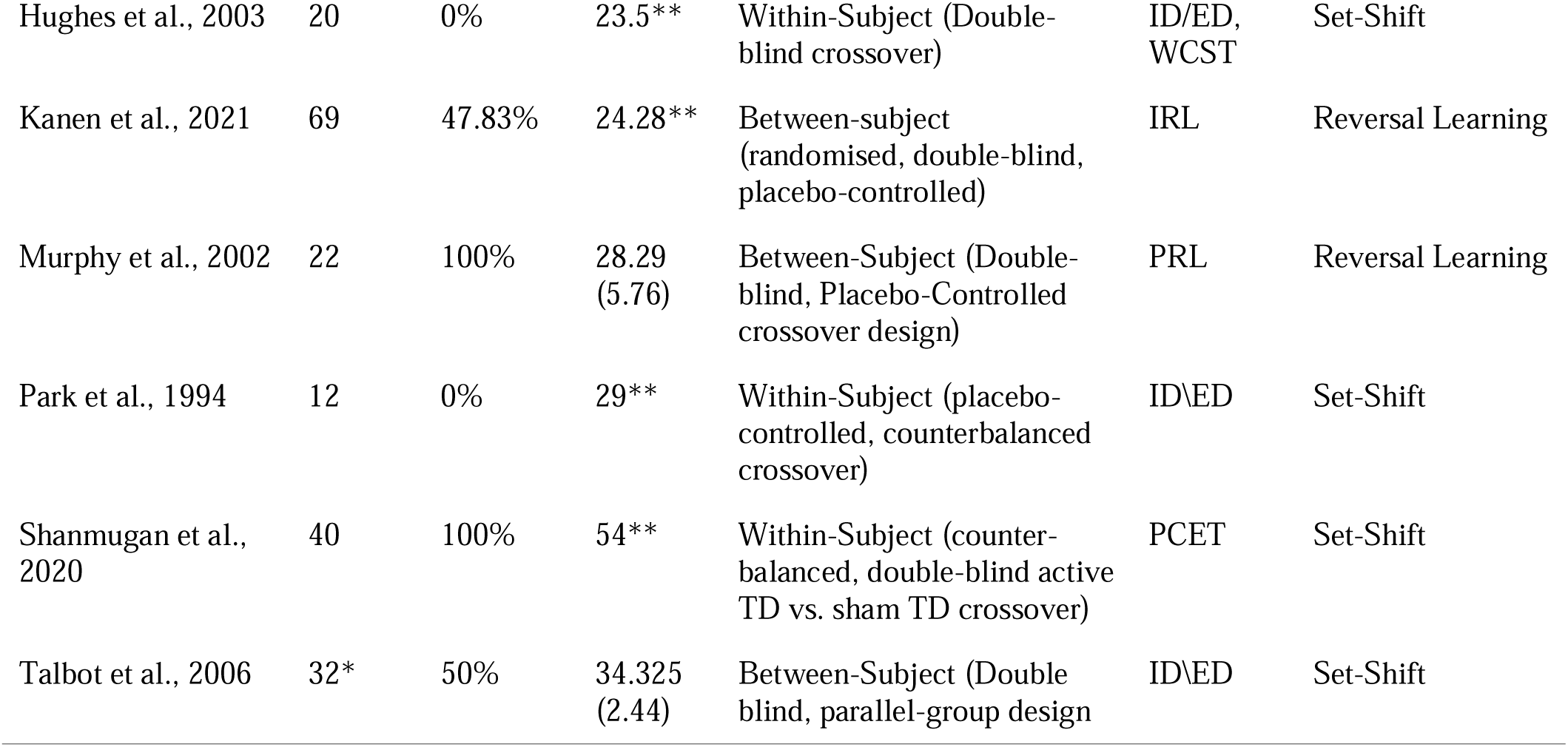
This table presents demographic and experimental details from the studies investigating the effects of ATD on performance in Cognitive flexibility tasks (Analysis A). including sample size, gender distribution, average age, study design, and cognitive task. * Talbot et al. (2006) had two relevant effect sizes - one with 32 participants and the other with 21 participants. Demographic data is given only regarding the initial sample. ** Some papers did not include the participants’ age SD. *** Abbreviations for cognitive tasks are as follows: WCST - Wisconsin Card Sorting Task, PRL - Probabilistic Reversal Learning, PCET - Penn Conditional Exclusion Test, ID/ED - Intra-Dimensional / Extra-Dimensional Set Shift, IRL - Instrumental Reversal Learning..

**Table 2.**
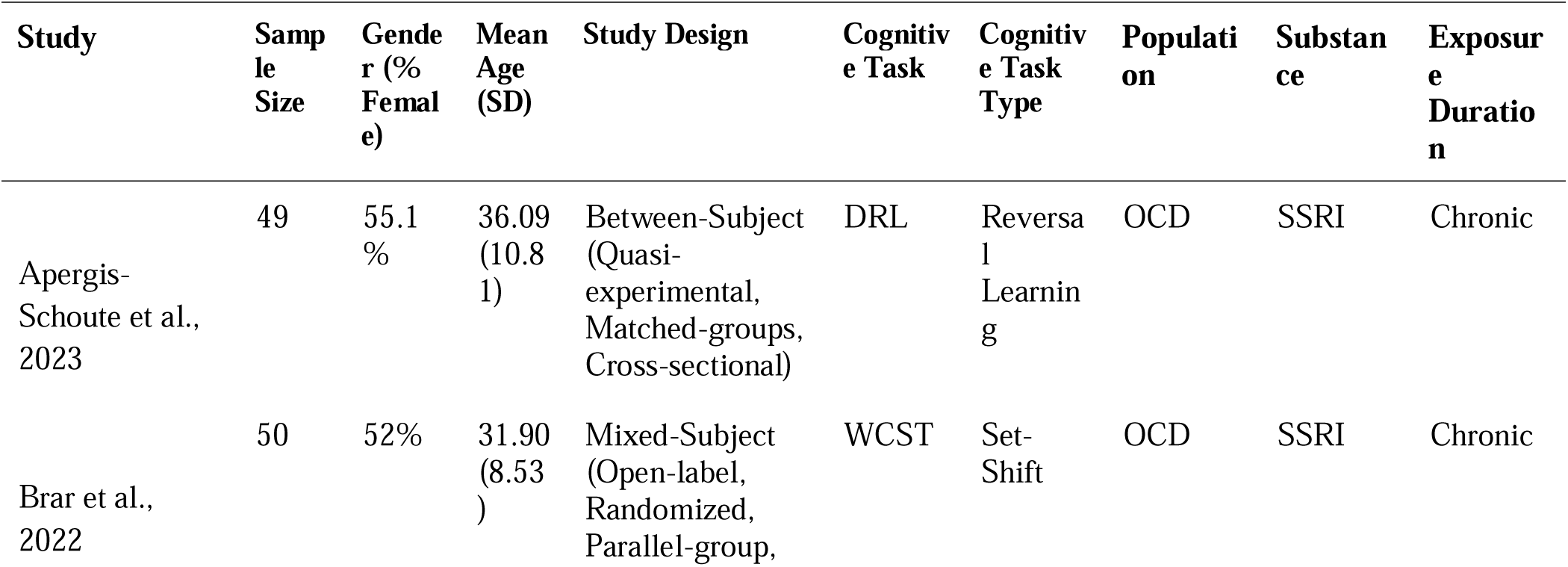
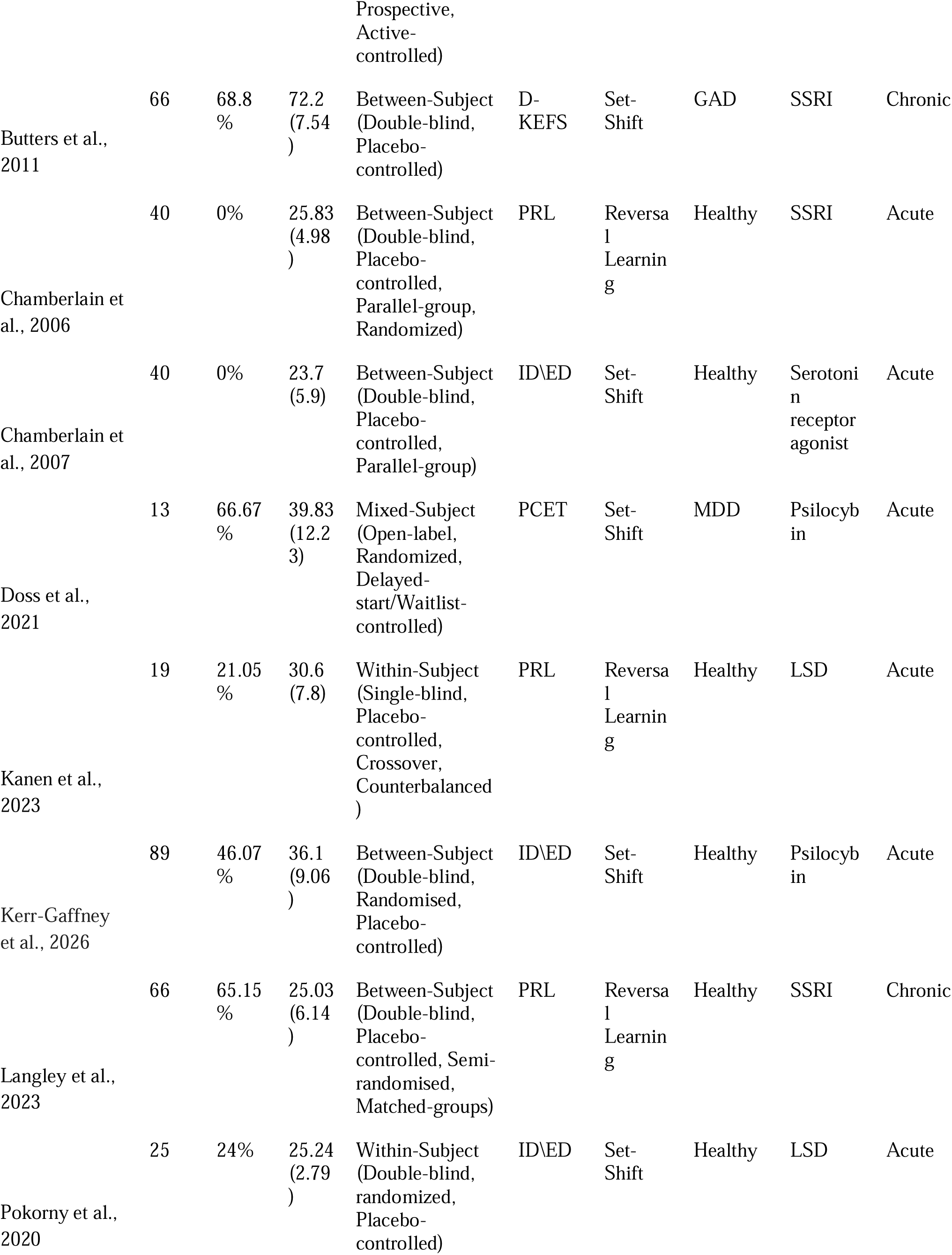
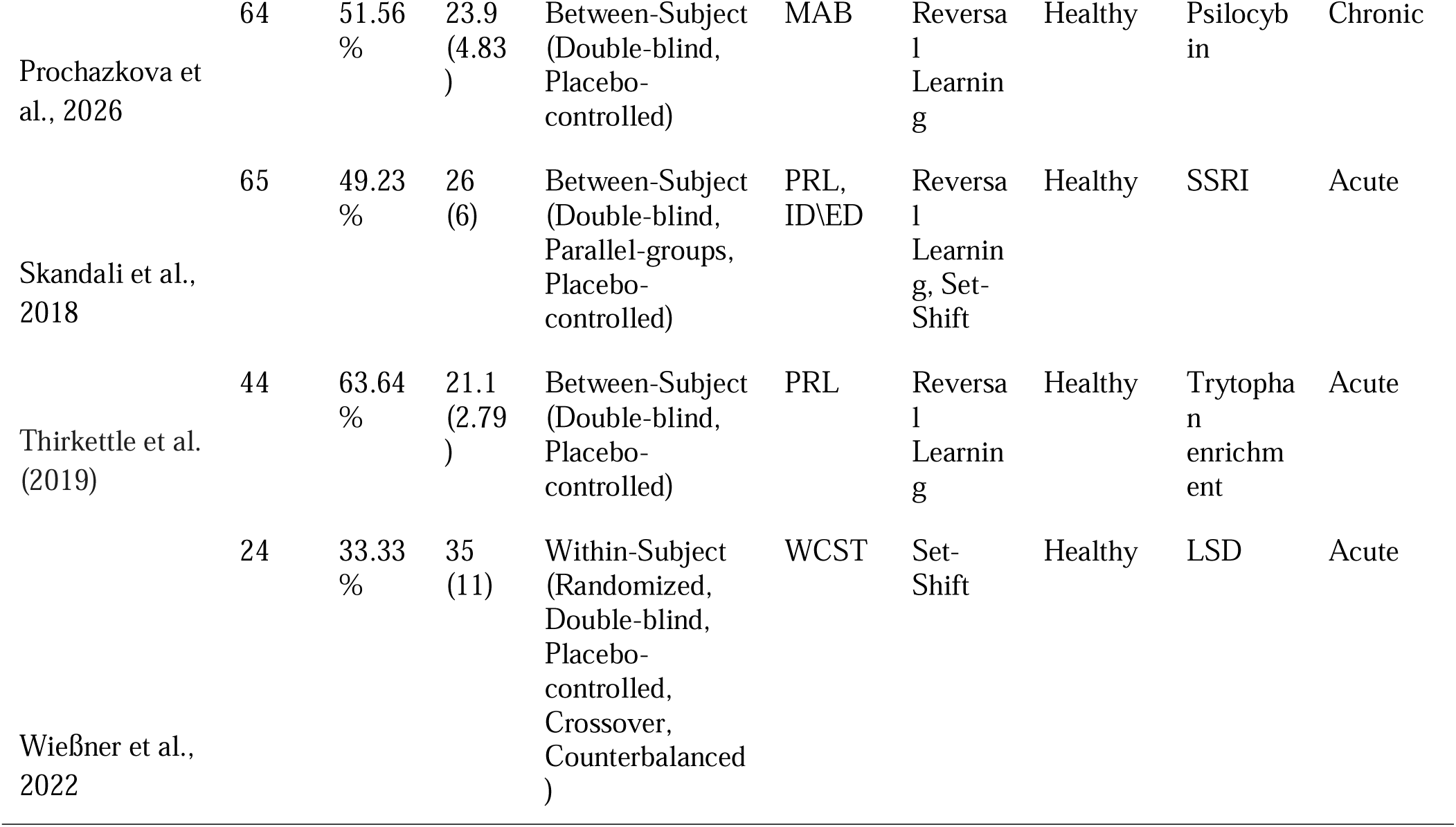
This table presents demographic and experimental details from the studies investigating the effects of serotonin elevation manipulations on performance in Cognitive flexibility tasks (Analysis B), including sample size, gender distribution, average age, study design, cognitive task, Population (Psychopathology) and Substance class used. * Abbreviations for cognitive tasks are as follows: WCST - Wisconsin Card Sorting Task, PRL - Probabilistic Reversal Learning, PCET - Penn Conditional Exclusion Test, ID/ED - Intra-Dimensional / Extra-Dimensional Set Shift, MAB - Multi-Armed Bandit Task, DRL - Deterministic Reversal Learning, D-KEFS - Delis–Kaplan Executive Function System. ** Abbreviations for Population are as follows: OCD - Obsessive-Compulsive Disorder, GAD - Generalized Anxiety Disorder, MDD - Major Depressive Disorder.

| Study | Sample Size | Gender (% Female) | Mean Age (SD) | Study Design | Cognitive Task | Cognitive Task Type | Population | Substance | Exposure Duration |
| --- | --- | --- | --- | --- | --- | --- | --- | --- | --- |
| Apergis-Schoute et al., 2023 | 49 | 55.1 % | 36.09 (10.81) | Between-Subject (Quasi-experimental, Matched-groups, Cross-sectional) | DRL | Reversal Learning | OCD | SSRI | Chronic |
| Brar et al., 2022 | 50 | 52% | 31.90 (8.53) | Mixed-Subject (Open-label, Randomized, Parallel-group, | WCST | Set-Shift | OCD | SSRI | Chronic |

SEROTONIN AND PERSEVERATIVE THINKING: META-ANALYSIS
|  |  |  |  | Prospective,<br>Active-controlled) |  |  |  |  |  |
| --- | --- | --- | --- | --- | --- | --- | --- | --- | --- |
| Butters et al.,<br>2011 | 66 | 68.8<br>% | 72.2<br>(7.54<br>) | Between-Subject<br>(Double-blind,<br>Placebo-controlled) | D-KEFS | Set-Shift | GAD | SSRI | Chronic |
| Chamberlain et<br>al., 2006 | 40 | 0% | 25.83<br>(4.98<br>) | Between-Subject<br>(Double-blind,<br>Placebo-controlled,<br>Parallel-group,<br>Randomized) | PRL | Reversa<br>l<br>Learnin<br>g | Healthy | SSRI | Acute |
| Chamberlain et<br>al., 2007 | 40 | 0% | 23.7<br>(5.9) | Between-Subject<br>(Double-blind,<br>Placebo-controlled,<br>Parallel-group) | ID\ED | Set-Shift | Healthy | Serotoni<br>n<br>receptor<br>agonist | Acute |
| Doss et al.,<br>2021 | 13 | 66.67<br>% | 39.83<br>(12.2<br>3) | Mixed-Subject<br>(Open-label,<br>Randomized,<br>Delayed-<br>start/Waitlist-<br>controlled) | PCET | Set-Shift | MDD | Psilocyb<br>in | Acute |
| Kanen et al.,<br>2023 | 19 | 21.05<br>% | 30.6<br>(7.8) | Within-Subject<br>(Single-blind,<br>Placebo-controlled,<br>Crossover,<br>Counterbalanced<br>) | PRL | Reversa<br>l<br>Learnin<br>g | Healthy | LSD | Acute |
| Kerr-Gaffney<br>et al., 2026 | 89 | 46.07<br>% | 36.1<br>(9.06<br>) | Between-Subject<br>(Double-blind,<br>Randomised,<br>Placebo-controlled) | ID\ED | Set-Shift | Healthy | Psilocyb<br>in | Acute |
| Langley et al.,<br>2023 | 66 | 65.15<br>% | 25.03<br>(6.14<br>) | Between-Subject<br>(Double-blind,<br>Placebo-controlled, Semi-<br>randomised,<br>Matched-groups) | PRL | Reversa<br>l<br>Learnin<br>g | Healthy | SSRI | Chronic |
| Pokorny et al.,<br>2020 | 25 | 24% | 25.24<br>(2.79<br>) | Within-Subject<br>(Double-blind,<br>randomized,<br>Placebo-controlled) | ID\ED | Set-Shift | Healthy | LSD | Acute |

Table 2. This table presents demographic and experimental details from the studies investigating the effects of serotonin elevation manipulations on performance in Cognitive flexibility tasks (Analysis B), including sample size, gender distribution, average age, study design, cognitive task, Population (Psychopathology) and Substance class used.
|  |  |  |  |  |  |  |  |  |  |
| --- | --- | --- | --- | --- | --- | --- | --- | --- | --- |
| Prochazkova et al., 2026 | 64 | 51.56 % | 23.9 (4.83) | Between-Subject (Double-blind, Placebo-controlled) | MAB | Reversal Learning | Healthy | Psilocybin | Chronic |
| Skandali et al., 2018 | 65 | 49.23 % | 26 (6) | Between-Subject (Double-blind, Parallel-groups, Placebo-controlled) | PRL, ID\ED | Reversal Learning, Set-Shift | Healthy | SSRI | Acute |
| Thirkettle et al. (2019) | 44 | 63.64 % | 21.1 (2.79) | Between-Subject (Double-blind, Placebo-controlled) | PRL | Reversal Learning | Healthy | Tryptophan enrichment | Acute |
| Wießner et al., 2022 | 24 | 33.33 % | 35 (11) | Within-Subject (Randomized, Double-blind, Placebo-controlled, Crossover, Counterbalanced) | WCST | Set-Shift | Healthy | LSD | Acute |

**Table 3.**
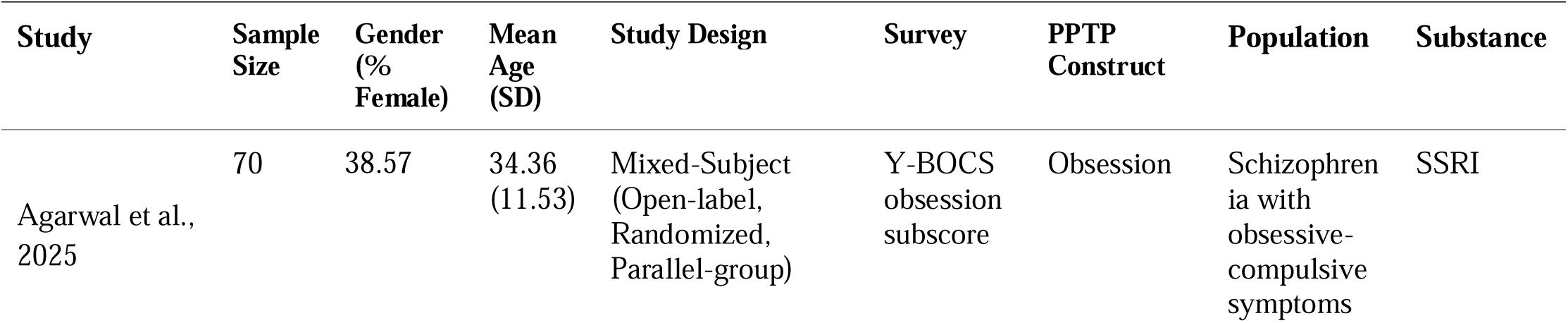
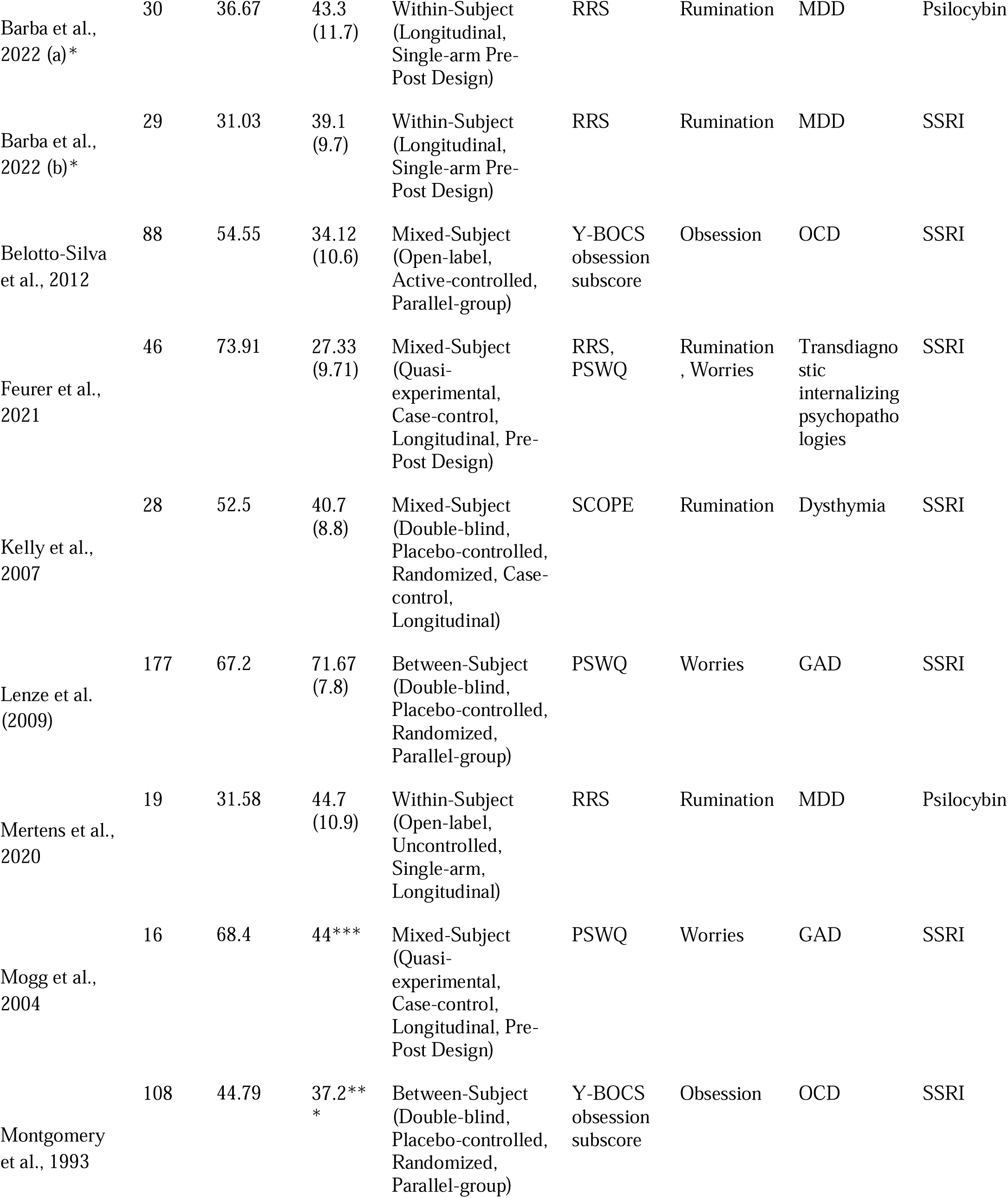
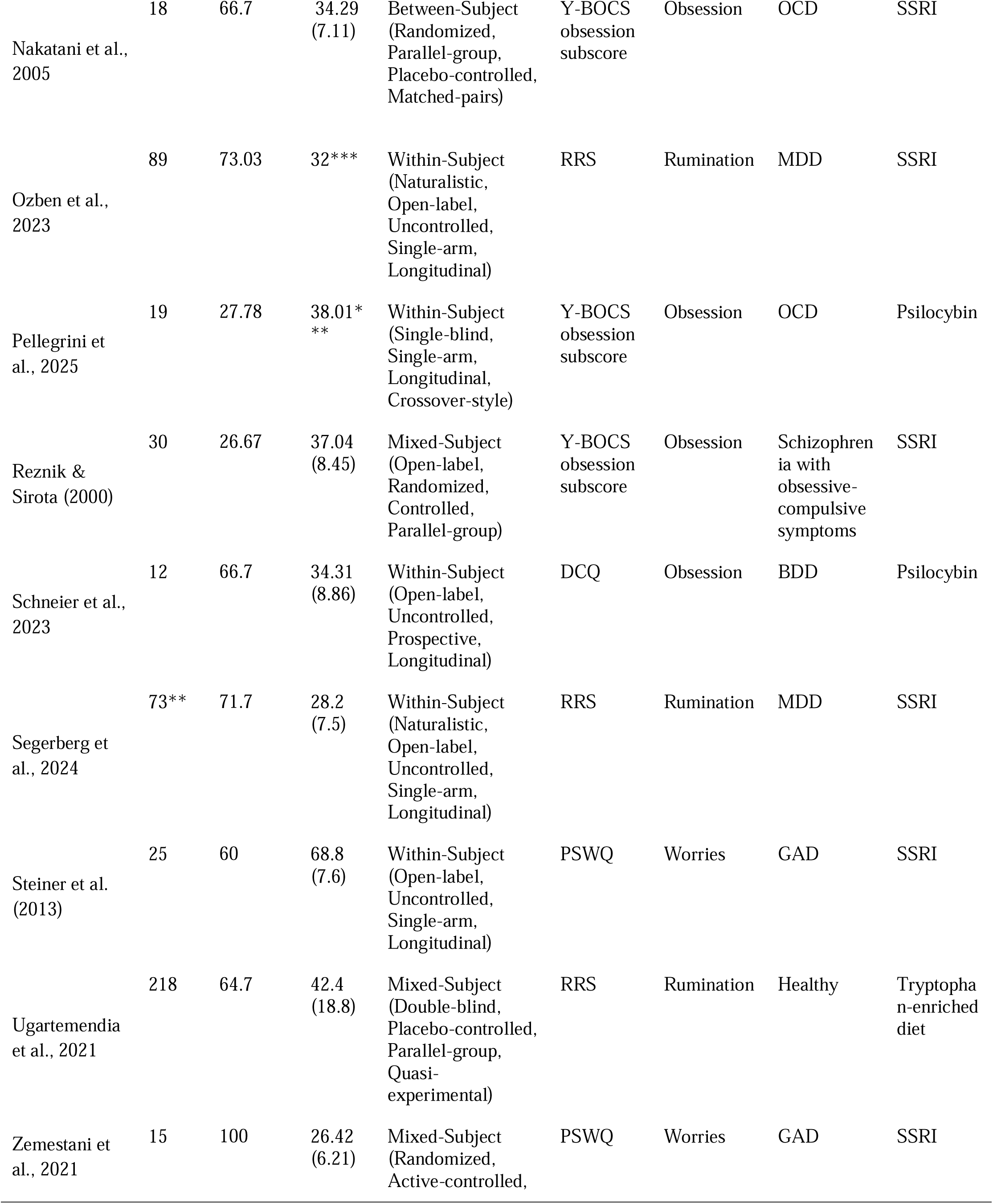
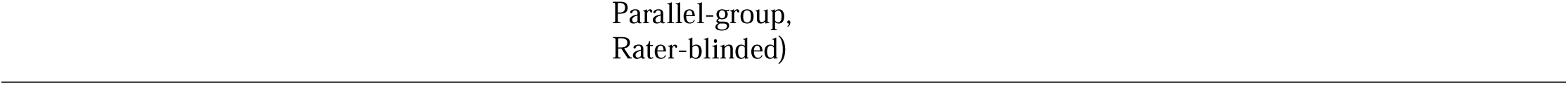
This table presents demographic and experimental details from the studies investigating the effects of serotonin elevation manipulations on PPTP questionnaires scores (Analysis B), including sample size, gender distribution, average age, study design, name of questionnaire, perseverative thinking pattern construct, Population (Psychopathology) and Substance class used. * Each treatment group (Psilocybin and SSRI) in Barba et al. (2022) was analyzed as a within-subject experiment standing for itself. ** Pre-treatment sample size in Segerberg et al. (2024) was 92, while in week 12 (the endpoint) it was 73. *** Some papers did not include the participants’ age SD. **** Abbreviations for pathological perseverative thinking patterns scales are as follows: RRS - Ruminative Response Scale, PSWQ - Penn State Worry Questionnaire, Y-BOCS - Yale-Brown Obsessive Compulsive Scale, DCQ - Dysmorphic Concerns Questionnaire, SCOPE - System of Coping Profile Endorsements. ***** Abbreviations for Population are as follows: OCD - Obsessive-Compulsive Disorder, GAD - Generalized Anxiety Disorder, MDD - Major Depressive Disorder, BDD - Body Dismorphic Disorder.

### Recorded Measures and Data Extraction

The recorded variables included general information, such as the study’s authors, year of publication, number of subjects, Cognitive task \ PT surveys and demographic variables, including age, sex, and serotonin manipulation dosage (see Table 1, 2 and 3). For the calculation of effect sizes, means and standard deviations (SD) were collected, of (1) perseverative errors, (2) number of completed categories, (3) total score of PPTP questionnaire. If this information was not available in the publication or provided by the authors, effect sizes were estimated using one of the following methods: (1) by extracting mean and SD from published figures (using PlotDigitizer software, available at http://plotdigitizer.sourceforge.net), (2) based on F values, or (3) based on p values.

#### Measures for cognitive flexibility (Analyses A & B)

● **Wisconsin Card Sorting Task (WCST):** Assesses the ability to shift cognitive strategies in response to changing environmental contingencies (Berg, 1948).
● **Probabilistic Reversal Learning (PRL):** Evaluates cognitive flexibility and reward processing by requiring participants to map changing stimulus-reward contingencies in the presence of misleading, probabilistic feedback (Cools et al., 2002).
● **Penn Conditional Exclusion Test (PCET):** Assesses executive functioning and abstract concept formation through a computerized sorting paradigm that requires participants to deduce changing categorization rules based on trial feedback (Kurtz et al., 2004).
● **Intra-Dimensional / Extra-Dimensional Set Shift (ID/ED):** Disentangles rule-learning within a specific stimulus dimension from the higher-order shifting of attention across different stimulus dimensions (Dowens et al., 1989).
● **Multi-Armed Bandit Task (MAB):** Captures cognitive flexibility by assessing how effectively participants update their choices and switch between different reward arms as payout values drift over time. This dynamic environment allows researchers to quantify flexible behavior through standardized metrics of exploratory switching and sequential decision-making (Daw et al., 2006; Mekern et al., 2019).
● **Deterministic reversal learning (DRL):** Assesses rapid rule-shifting and behavioral adaptation under high executive load and time pressure, where a failure to switch response strategies following a 100% predictable reversal in feedback contingencies is indexed by repeated errors (errors followed by another error) and a higher number of trials required to re-attain the learning criterion (Apergis-Schoute et al., 2024).
● **Delis–Kaplan Executive Function System (D-KEFS)**: Assesses cognitive flexibility primarily through subtests like Trail Making and Color-Word Interference, which require participants to dynamically switch between competing rules or visual sets. By measuring an individual’s ability to inhibit automatic responses and shift focus under changing conditions, it provides a standardized metric of set-shifting capabilities (Delis et al., 2001).
● **Instrumental Reversal Learning (IRL):** Measures behavioral adaptation, translational rule shifting, and reinforcement learning by requiring participants to map changing stimulus-response contingencies under high executive load, using serial deterministic reversals paired with varying valences of salient visual and auditory feedback (reward, punishment, and neutral conditions) across complex bimanual motor mappings (Apergis-Schoute et al., 2020).

#### Measures for pathological perseverative thinking patterns (Analysis C)

● **Ruminative Response Scale (RRS):** Evaluates the tendency to engage in different ruminative thinking patterns (Nolen-Hoeksema & Morrow, 1991).
● **Penn State Worry Questionnaire (PSWQ):** Quantifies the generality, intensity, and uncontrollability of clinical and non-clinical chronic worry (Meyer et al., 1990).
● **Yale-Brown Obsessive Compulsive Scale (Y-BOCS) - Obsession Subscale:** Measures the severity, frequency, duration, and associated distress of intrusive, persistent thoughts independently of compulsive behaviors (Goodman et al., 1989).
● **Dysmorphic Concerns Questionnaire (DCQ):** Screens for overvalued ideas and obsessive worry regarding perceived physical flaws or defects in bodily appearance (Oosthuizen et al., 1998).
● **System of Coping Profile Endorsements (SCOPE) - Rumination Subscale:** Evaluates the specific tendency to cope with environmental or emotional stressors through passive, repetitive cognitive focus on the stressor (Matheson & Anisman, 2003).

## Statistical Analyses

All statistical analyses were conducted in RStudio (version 2026.05.0-218) utilizing the metafor (Viechtbauer, 2010) and clubSandwich (Pustejovsky, 2020) packages. Effect sizes were calculated as Hedges’ g to correct for small-sample bias, with interpretive thresholds aligned with Cohen’s d conventions. In this analysis, negative effect sizes indicate an improvement in cognitive flexibility performance. Specifically, this corresponds to a reduction in perseverative errors, an increased number of categories achieved, or lower scores on measures of perseverative thinking. Parameter estimation for all models was performed using restricted maximum likelihood (REML), and heterogeneity was evaluated using the Q-statistic and residual variance (τ^2). For primary studies reporting only medians and interquartile ranges, sample means and standard deviations were estimated using the mathematical approximations established by Wan et al. (2014).

### Handling Statistical Dependency

Because several included studies reported multiple outcomes from the same participant cohorts (e.g., multiple timepoints, overlapping subscales, or distinct cognitive measures), a two-pronged approach was employed to appropriately model statistical dependency without sacrificing the resolution needed for moderator analyses.

First, when multiple effect sizes from a single study represented variations that were not of theoretical interest as independent moderators (e.g., multiple measurement timepoints), these dependent effect sizes were aggregated into a single composite effect size prior to global pooling. This targeted aggregation was utilized primarily in Analyses A and B. Following Borenstein et al. (2009), the aggregated variance was computed by adjusting for the intra-study correlation (r) among outcomes to avoid artificially inflating precision. Based on conservative recommendations in psychological meta-analyses, this correlation was set a priori to r = 0.50.

Second, when multiple effect sizes within a study represented distinct categories of a planned within-study moderator (such as the distinct phenomenological constructs of rumination, worry, and obsession in Analysis C), prior aggregation was not performed. Instead, these effect sizes were retained as separate data points.

To robustly handle both these disaggregated data points and any residual dependencies across all analyses, the global meta-analyses were executed using a multilevel random-effects framework (via the rma.mv function), nesting individual effect sizes within their respective study identifiers. Furthermore, cluster-robust variance estimation (RVE) utilizing the “CR2” estimator was applied to all models. This technique robustly adjusts standard errors and confidence intervals to account for correlated sampling errors within studies, ensuring that Type I error rates are controlled even when multiple effect sizes are drawn from the same participant sample.

### Moderator Analyses

Three separate multilevel meta-regression models were performed to address the primary research questions, incorporating the following a priori moderators:

● Analysis A (Effects of ATD on cognitive flexibility): Biological sex, mean age, and cognitive task type.
● Analysis B (Effects of serotonin elevation on cognitive flexibility): Substance class (SSRI, LSD, Psilocybin and Tryptophan enrichment), exposure duration (acute vs. chronic), biological sex, and mean age.
● Analysis C (Effects of serotonin elevation on PPTP): Phenomenological construct (rumination, worries, obsessions), specific serotonin manipulation, biological sex, and mean age.

In Analysis C, to prevent severe multicollinearity and ensure the stability of parameter estimates, exposure duration (acute vs. chronic) was omitted from the moderator analysis. This decision was made because exposure duration perfectly covaried with substance type in our dataset, rendering the independent effects of acute versus chronic administration mathematically indistinguishable from the specific serotonin manipulation

### Publication Bias

Potential publication bias and small-study effects were evaluated via visual inspection of funnel plots and quantitatively assessed using Egger’s regression test for funnel plot asymmetry, with statistical significance defined as p <.05.

## Results

### Analysis A: The Effect of ATD on Cognitive Flexibility

#### General Effect

The estimated intercept of the overall multilevel random-effects model (without moderators) was 0.1546 (*SE* = 0.1169, *z* = 1.3227, *p* = 0.1859, 95% CI [-0.0745, 0.3837]). When adjusting for correlated sampling errors using cluster-robust variance estimation (RVE), the overall effect remained non-significant (*t* = 1.32, *df* = 7.23, *p* = 0.2260). This indicates that, across all studies, a transient decrease in central serotonin via acute tryptophan depletion (ATD) did not yield a statistically significant overall effect on objective performance in cognitive flexibility measures.

#### Effects of Cognitive Task Type, Biological Sex, and Age

The full multilevel meta-regression model exploratorily evaluated whether the pooled effect was moderated by cognitive task type (set-shifting vs. reversal learning), biological sex (proportion of female participants), and mean age. Utilizing the CR2 robust standard errors to control for Type I error inflation, none of the examined covariates reached strict statistical significance:

● Cognitive task type did not significantly moderate the effect (Estimate =-0.5830, CR2 *SE* = 0.3889, *t* =-1.50, *p* = 0.2322), indicating no substantive difference in effect sizes between cue-based set-shifting and feedback-driven reversal learning paradigms.
● The effect of biological sex was not statistically significant after robust adjustments (Estimate =-1.2812, CR2 *SE* = 0.5943, *t* =-2.16, *p* = 0.1407), indicating that the proportion of female participants in a sample did not reliably predict the magnitude of the effect.
● The effect of mean age demonstrated a marginal trend (Estimate = 0.0575, CR2 SE = 0.0224, *t* = 2.56, *p* = 0.0757). This suggests a potential, weak positive association where older age might be associated with stronger negative effect of ATD on cognitive flexibility, though this did not reach the p < 0.05 threshold for significance.

### Model Fit and Heterogeneity

The baseline model revealed moderate heterogeneity, with an estimated τ² value of 0.0612 (*SE* = 0.0560). The I² statistic was calculated to be 53.33%, indicating that roughly half of the variability in effect sizes was due to true heterogeneity across studies rather than pure sampling error. In the meta-regression model, the omnibus test of moderators (*QM*(3) = 5.7144, *p* = 0.1264) indicated that the inclusion of cognitive task type, sex, and age collectively did not explain a statistically significant proportion of the variability in effect sizes. Additionally, the test for residual heterogeneity was non-significant (*QE*(6) = 8.0726, *p* = 0.2328), suggesting that the remaining variance in the model was within the bounds of expected sampling error.

### Publication Bias

To assess publication bias and small-study effects, Egger’s regression test was conducted. The test revealed a non-significant result (*t* =-0.9186, *df* = 8, *p* = 0.3852), indicating a lack of significant funnel plot asymmetry and suggesting that the results are not substantially driven by small-study publication bias.

We further performed influence analysis. The influence analysis, which identifies individual studies that might disproportionately skew the overall model parameters, did not detect any disproportionately influential studies within the dataset.

**Figure 3A.**
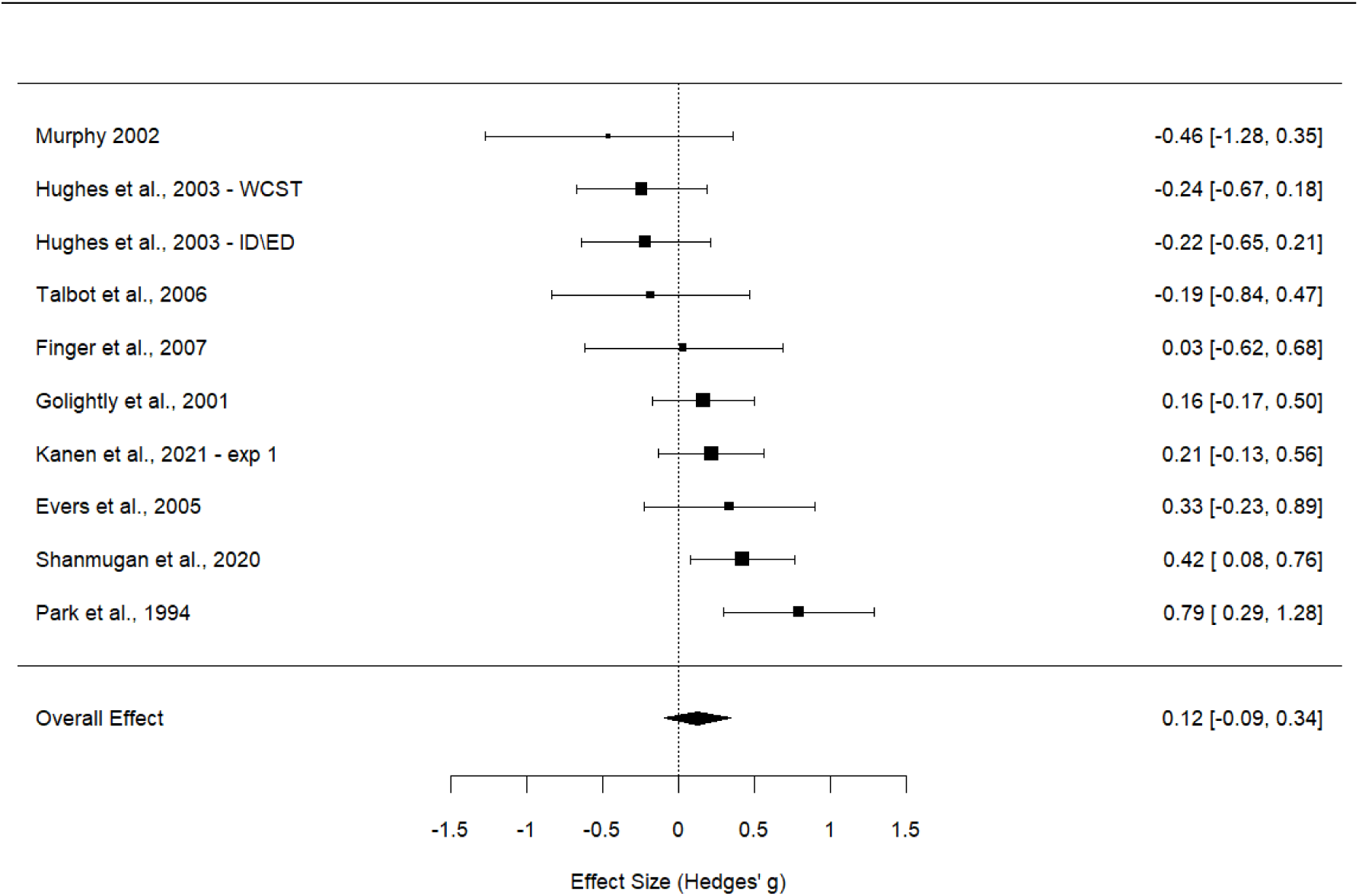
A forest plot of the effect sizes for each study included in Analysis A (acute tryptophan depletion). Points show effect sizes (Hedges’ g) with 95% confidence intervals. The diamond indicates the overall pooled effect.

**Figure 3B.**
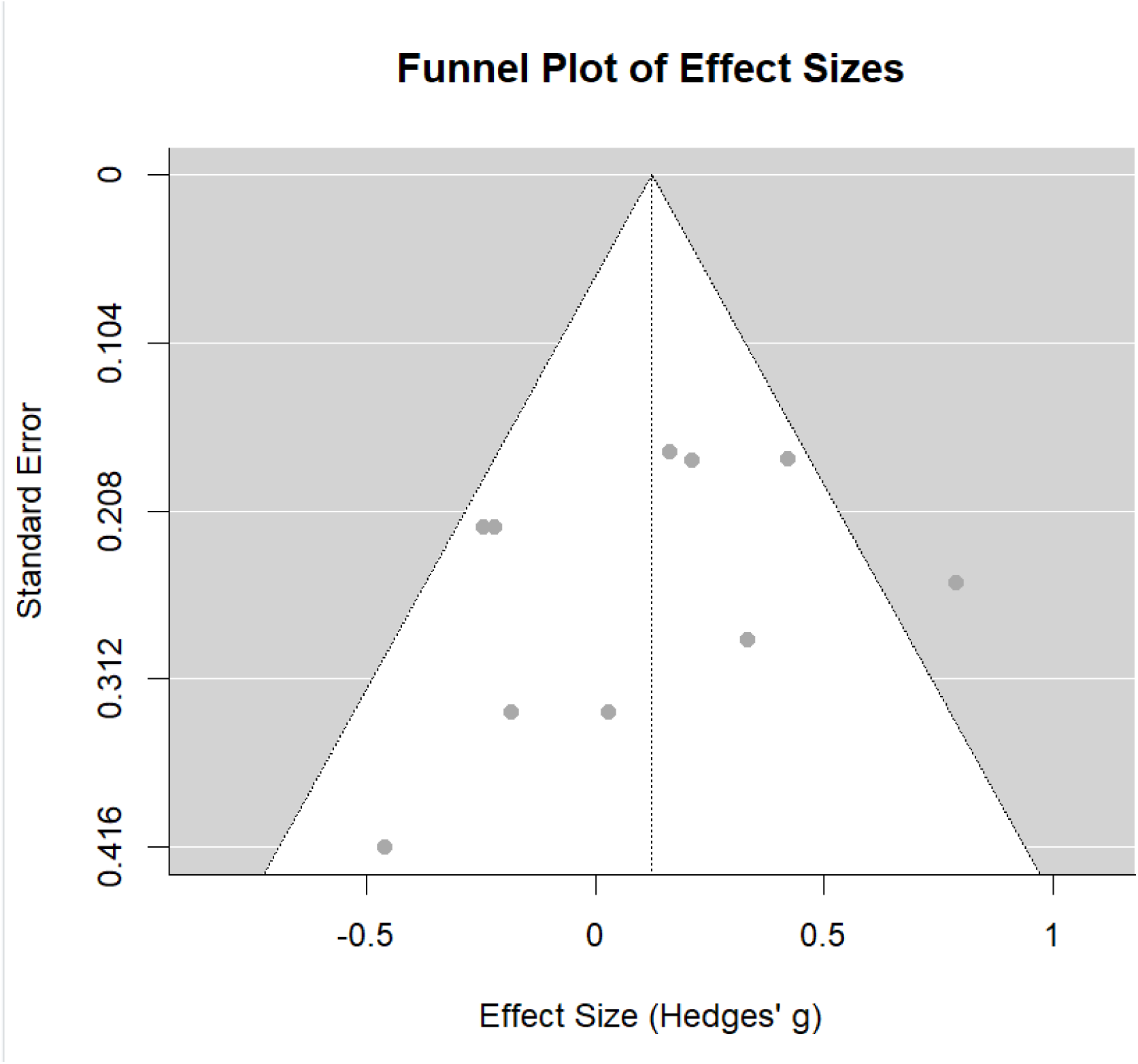
A funnel plot of the effect sizes for studies in Analysis A to visually assess potential publication bias. Individual study effect sizes (Hedges’ g) are plotted against their standard errors. The vertical dotted line represents the overall estimated effect, and the diagonal dashed lines indicate the pseudo-95% confidence limits. The symmetrical distribution of the data points around the overall effect estimate suggests an absence of substantial small-study publication bias.

### Analysis B: The Effect of Serotonin Elevation on Cognitive Flexibility

#### General Effect

The estimated intercept of the overall multilevel random-effects model (without moderators) was-0.0729 (*SE* = 0.1473, *z* =-0.4951, *p* = 0.6205, 95% CI [-0.3617, 0.2158]). When adjusting for correlated sampling errors using cluster-robust variance estimation (RVE), the overall effect remained statistically non-significant (*t* =-0.496, *df* = 12.9, *p* = 0.6280). This indicates that, across all included studies, an elevation of serotonergic activity did not yield a statistically significant overall effect on objective cognitive flexibility measures.

#### Effects of Substance Class, Exposure Timeline, Biological Sex, and Age

The full multilevel meta-regression model exploratorily evaluated whether the pooled effect was moderated by specific substance class, exposure timeline parameters (acute vs. chronic schedules), biological sex (proportion of female participants), and mean age. Utilizing the CR2 robust standard errors to control for Type I error inflation, the covariates did not yield robust statistically significant effects:

● Comparing specific serotonin manipulations to the reference group (SSRIs), neither LSD (Estimate = 0.5685, CR2 *SE* = 0.3804, *t* = 1.495, *p* = 0.2050), psilocybin (Estimate =-0.3991, CR2 *SE* = 0.4534, *t* =-0.880, *p* = 0.4700), nor tryptophan enrichment (Estimate =-0.5863, CR2 *SE* = 0.6304, *t* =-0.930, *p* = 0.4630) significantly moderated the effect size. This indicates no distinct advantage or variation in cognitive flexibility outcomes based on the specific pharmacological agent when compared to standard SSRI administration.
● The exposure timeline parameter did not significantly moderate the effect (Estimate =-0.0828, CR2 *SE* = 0.5459, *t* =-0.152, *p* = 0.8930), suggesting that acute versus chronic administration schedules do not yield substantively different effect magnitudes.
● The effect of biological sex was not statistically significant (Estimate = 0.1787, CR2 *SE* = 1.0418, *t* = 0.172, *p* = 0.8750), indicating that the proportion of female participants in a sample did not influence the cognitive flexibility outcomes.
● The effect of mean age was also not statistically significant (Estimate =-0.0180, CR2 *SE* = 0.0194, *t* =-0.924, *p* = 0.4550), demonstrating that variations in sample age did not meaningfully alter the overall effect.

#### Model Fit and Heterogeneity

The baseline model revealed substantial heterogeneity, with an estimated τ² value of 0.2365 (*SE* = 0.1129). The I² statistic was calculated to be 80.88%, indicating that the vast majority of the variability in effect sizes was due to true heterogeneity across studies rather than random sampling error. In the meta-regression model, the omnibus test of moderators (*QM*(6) = 8.6931, *p* = 0.1916) confirmed that the inclusion of substance class, exposure timeline, sex, and age collectively did not explain a statistically significant proportion of the variability in effect sizes. The test for residual heterogeneity remained highly significant (*QE*(8) = 29.9585, *p* = 0.0002), underscoring the ongoing presence of considerable unexplained variance across the studies even after accounting for the exploratory covariates.

### Publication Bias

To assess publication bias and small-study effects, Egger’s regression test was conducted. The test revealed a non-significant result (*t* = 0.0645, *df* = 13, *p* = 0.9496), indicating no evidence of funnel plot asymmetry. This suggests that the distribution of effect sizes is symmetrical and that the analysis is not substantially driven by small-study publication bias.

We further performed influence analysis, which did not detect any disproportionately influential outliers within the dataset, reflecting stability in the parameter estimation.

**Figure 4A.**
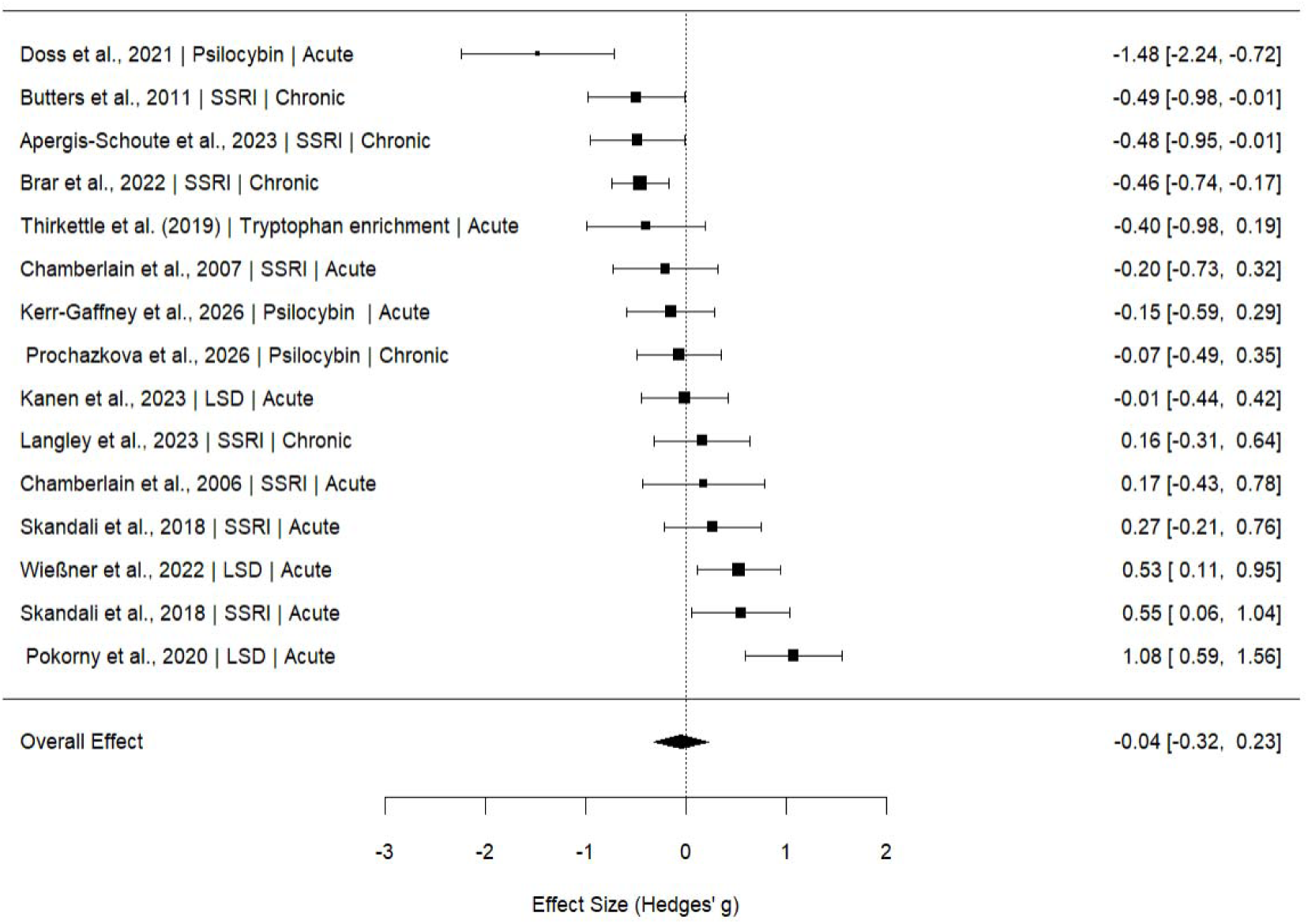
A forest plot of the effect sizes for each study included in Analysis B (serotonin elevation manipulations). Points show effect sizes (Hedges’ g) with 95% confidence intervals. The diamond indicates the overall pooled effect. Studies are annotated by their respective substance class (e.g., SSRI, Psilocybin, LSD, Tryptophan enrichment) and exposure timeline (Acute vs. Chronic).

**Figure 4B.**
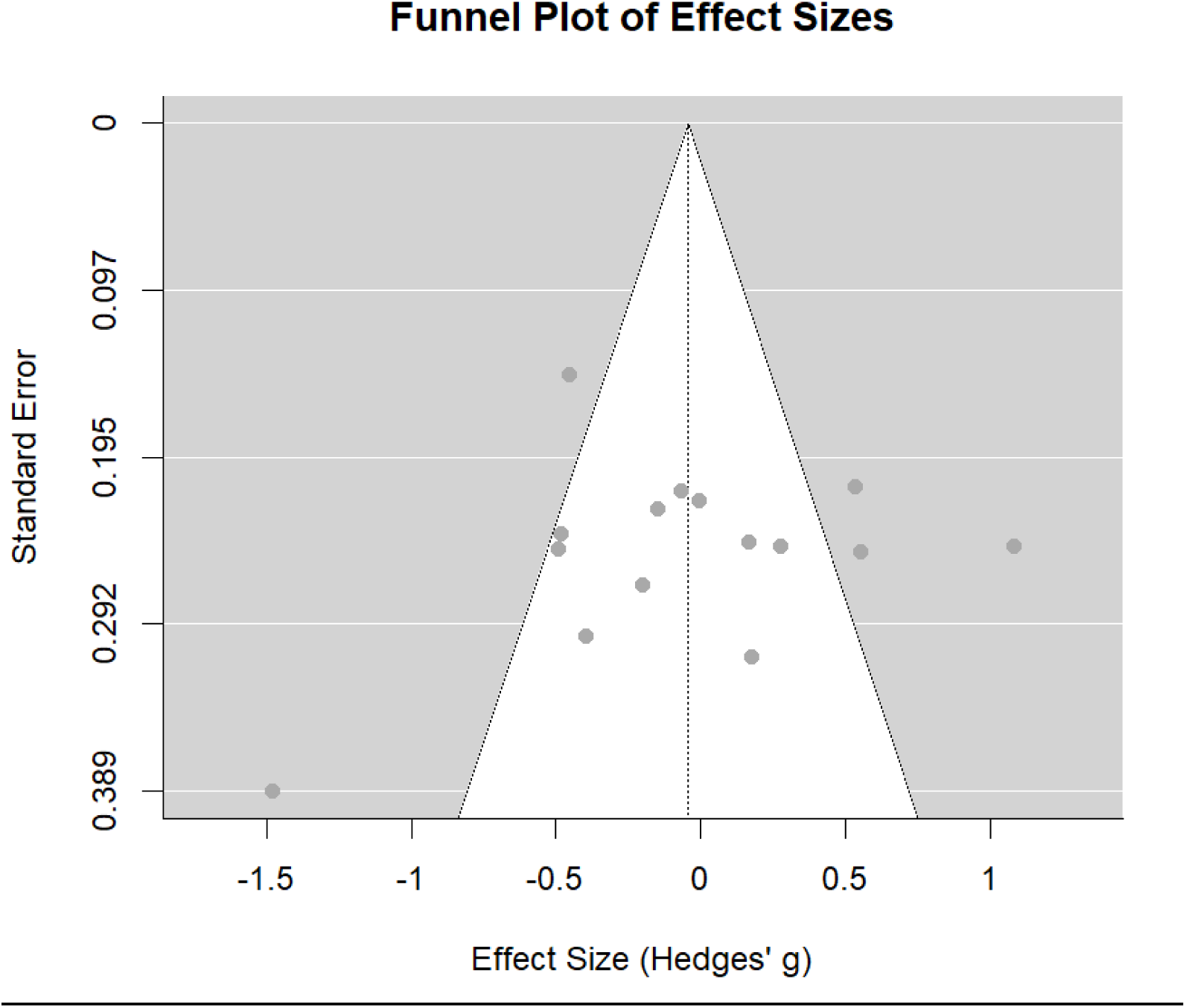
funnel plot of the effect sizes for studies in Analysis B to visually assess potential publication bias. Individual study effect sizes (Hedges’ g) are plotted against their standard errors. The vertical dotted line represents the overall estimated effect, and the diagonal dashed lines indicate the pseudo-95% confidence limits. The symmetrical distribution of the data points around the overall effect estimate suggests an absence of substantial small-study publication bias.

### Analysis C: The Effect of Serotonin Elevation on the Decrease in Perseverative Thinking

#### General Effect

The estimated intercept of the overall multilevel random-effects model (without moderators) was-0.5849 (*SE* = 0.0890, *z* =-6.5724, *p* < 0.0001, 95% CI [-0.7594,-0.4105]). When adjusting for correlated sampling errors using cluster-robust variance estimation (RVE), the overall effect remained highly significant (*t* =-6.55, *df* = 17, p < 0.001). This indicates that, across all studies, serotonin elevation was associated with a statistically significant, medium-to-large overall reduction in the measured phenomenological constructs (rumination, worries, and obsessions).

#### Effects of Phenomenological Construct, Substance, Sex, and Age

The full multilevel meta-regression model incorporated the phenomenological construct, specific serotonin manipulation (substance), biological sex, and mean age.

● The effect of biological sex (proportion of female participants) was statistically significant, with an estimated coefficient of-1.8552 (CR2 *SE* = 0.5218, *t* =-3.5551, *p* = 0.0144). Because the overall effect sizes are negative, this finding indicates that samples with a higher proportion of female participants exhibited a significantly larger reduction in symptoms.
● Regarding specific serotonin manipulations, using SSRIs as the reference group, the effect size for acute psilocybin interventions demonstrated a marginally significant trend toward greater symptom reduction (Estimate =-0.3219, CR2 *SE* = 0.1476, *t* = - 2.1811, *p* = 0.0809). This suggests a potential trend wherein chronic SSRI administration, while effective, yielded slightly smaller magnitudes of symptom reduction compared to acute psilocybin. Additionally, the single study utilizing a tryptophan-enriched diet was significantly different from the SSRI reference group (Estimate = 0.6776, CR2 *SE* = 0.1737, *t* = 3.9005, *p* = 0.0132), indicating a significantly smaller reduction in perseverative thinking compared to the SSRI baseline.
● Regarding the phenomenological construct, using obsessions as the reference group, rumination did not show a statistically significant difference (Estimate =-0.1552, CR2 *SE* = 0.1681, *t* =-0.9231, *p* = 0.3831). However, the worries construct showed a marginally significant positive coefficient (Estimate = 0.4247, CR2 *SE* = 0.1574, *t* = 2.6983, *p* = 0.0545), suggesting that symptom reduction for worries was potentially smaller in magnitude compared to obsessions.
● The effect of mean age was not statistically significant (Estimate =-0.0003, CR2 *SE* = 0.0051, *t* =-0.0536, *p* = 0.9610), indicating that sample age did not meaningfully moderate the effect size.

#### Model Fit and Heterogeneity

The baseline model revealed substantial residual heterogeneity, with an estimated τ² value of 0.1473 (*SE* = 0.0629), suggesting a high degree of unexplained variability among the effect sizes. The I² statistic was calculated to be 83.12%, indicating that most of the variability in effect sizes was due to true heterogeneity rather than sampling error. In the meta-regression model, the omnibus test of moderators (*QM*(6) = 17.6572, *p* = 0.0071) indicated that the inclusion of construct, substance, biological sex, and age collectively explained a significant proportion of the variability in effect sizes. However, the test for residual heterogeneity remained highly significant (*QE*(13) = 39.8133, *p* = 0.0001), confirming the presence of considerable unexplained variance across studies even after accounting for these moderators.

#### Publication Bias

To assess publication bias and small-study effects, Egger’s regression test was conducted. The test revealed a non-significant result (*t* =-1.7822, *df* = 18, *p* = 0.0916). While there is a slight trend, this indicates that there is no statistically significant funnel plot asymmetry, suggesting that our overall findings are not substantially driven by small-study publication bias.

We further performed influence analysis and Fail-safe N calculations. The influence analysis, which identifies individual studies that might disproportionately affect the overall model parameters, revealed that Feurer et al. (2021; RRS as dependent variable measure) was an influential study. To assess the impact of this outlier, we re-ran the full meta-regression after omitting this specific effect size. Reassuringly, the main effect of serotonin elevation on perseverative thinking remained highly significant, confirming the robustness of the overall symptom reduction. However, the omission of this study diluted the strength of some moderators: biological sex and the differences between SSRI and tryptophan enrichment shifted to marginally significant trends, while the variance between phenomenological constructs (worries versus obsessions) was no longer significant. The effect size for acute psilocybin interventions maintained a marginally significant trend toward greater symptom reduction, which slightly strengthened after the removal of the outlier (Estimate =-0.3567, CR2 *SE* = 0.1547, *t* =-2.305, *p* = 0.0696).

Complementarily, the Rosenthal Fail-safe N was calculated to be 1504, indicating that an additional 1,504 studies with null effects would be necessary to invalidate our significant overall finding (target significance level = 0.05). This exceptionally high Fail-safe N well exceeds the conventional tolerance threshold, demonstrating the strength and reliability of the observed symptom reductions and confirming that potential unpublished null studies would not easily negate the significant impact of these serotonin manipulations.

**Figure 5A.**
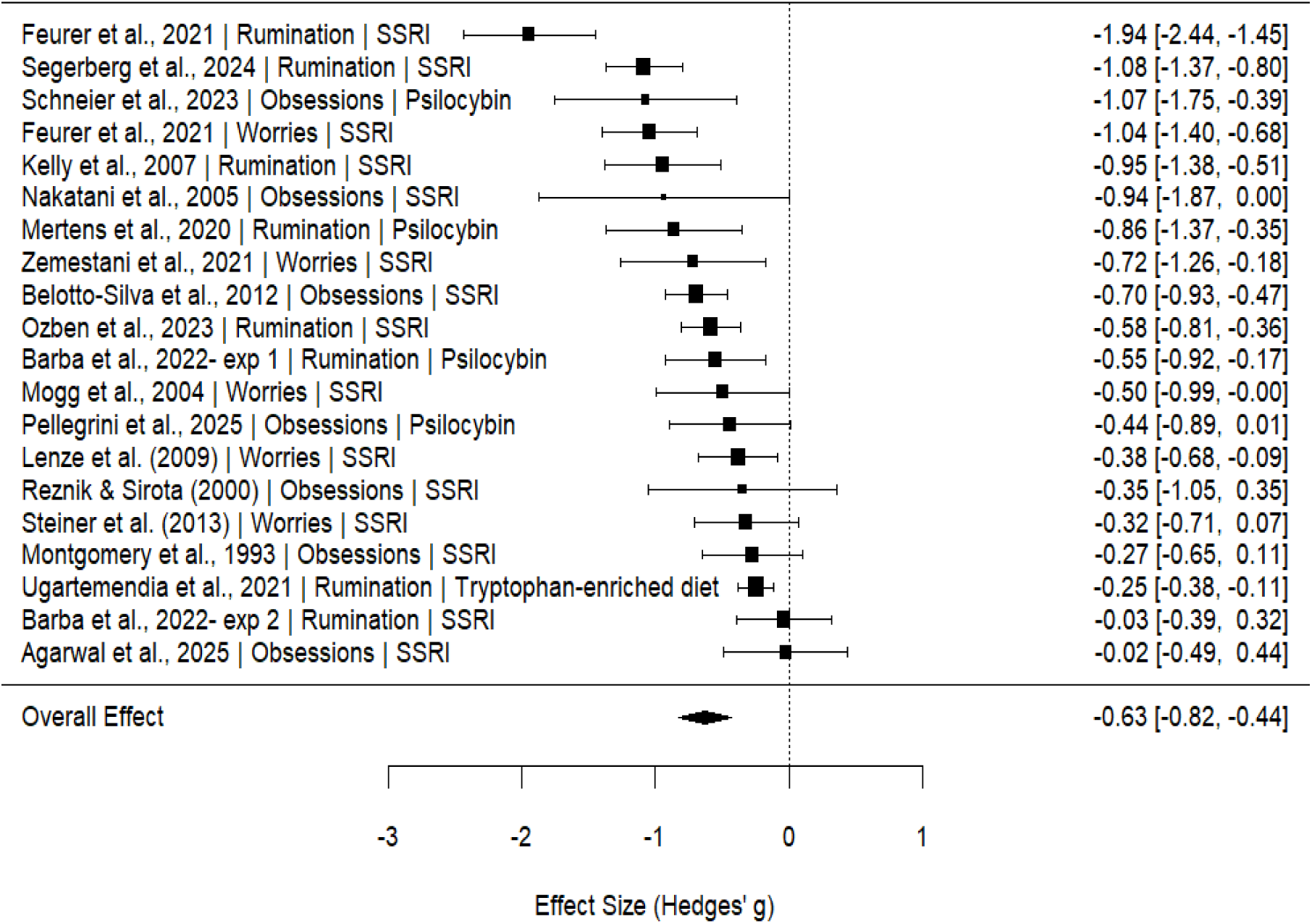
A forest plot of the effect sizes for each study included in Analysis C (the effect of serotonin elevation on perseverative thinking patterns). Points show effect sizes (Hedges’ g) with 95% confidence intervals. The diamond indicates the overall pooled effect. Studies are annotated by their respective phenomenological construct (Rumination, Worries, or Obsessions) and specific substance class (SSRI, Psilocybin, or Tryptophan-enriched diet).

**Figure 5B.**
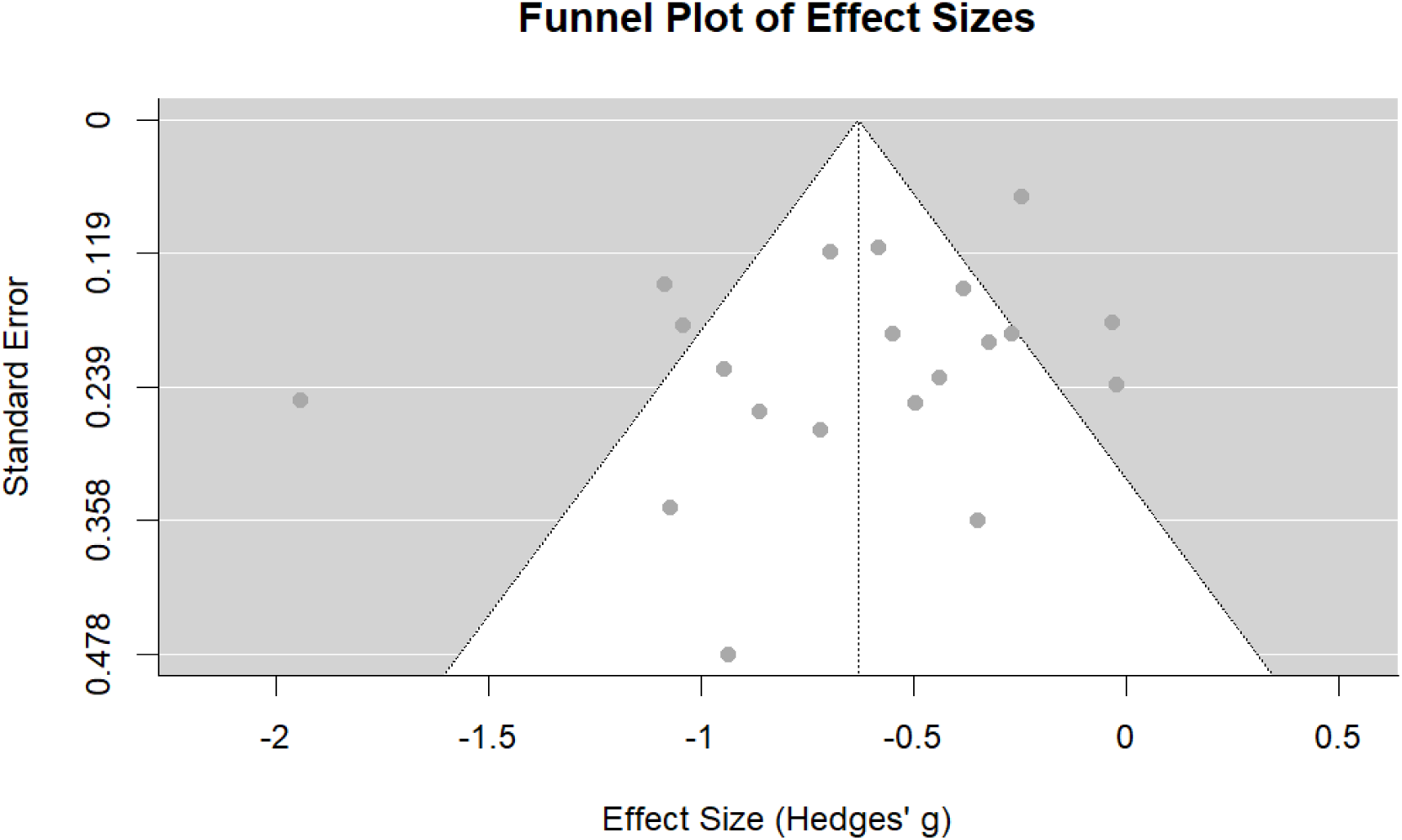
A funnel plot of the effect sizes for studies in Analysis C to visually assess potential publication bias. Individual study effect sizes (Hedges’ g) are plotted against their standard errors. The vertical dotted line represents the overall estimated effect, and the diagonal dashed lines indicate the pseudo-95% confidence limits. The plot includes all initial effect sizes prior to the removal of the identified outlier during the sensitivity analysis.

## Discussion

The present meta-analysis examined the effects of serotonergic manipulations on perseverative thinking patterns and cognitive flexibility. Across three complementary analyses, a clear dissociation emerged. Serotonin elevation was associated with a robust reduction in pathological perseverative thinking, including rumination, worries, and obsessions. However, acute tryptophan depletion did not significantly impair objective cognitive flexibility, and serotonin elevation through SSRIs and psychedelics did not significantly improve objective cognitive flexibility. Thus, contrary to the hypothesis that serotonergic effects on perseverative thinking would be primarily explained by improvements in cognitive flexibility, the current findings suggest that serotonin may influence repetitive negative thought more strongly than it influences performance on standard cognitive flexibility tasks. Serotonin has often been linked to behavioral adaptation, learning, affective regulation, and cognitive flexibility (Cools et al., 2008; Carhart-Harris & Nutt, 2017; Roberts et al., 2020; Dayan & Huys, 2009). Nevertheless, the current results suggest that this relationship may not be expressed as a broad, domain-general improvement in laboratory-based cognitive flexibility tasks. Rather, serotonergic interventions may influence the subjective, affective, and self-referential experience of thought. In other words, serotonin may help individuals disengage from repetitive negative thinking without necessarily improving their performance on objective set-shifting or reversal-learning tasks.

A central interpretation of these findings concerns the distinction between objective cognitive performance and subjective self-report perception of psychological functioning. Cognitive flexibility tasks typically assess performance under structured laboratory conditions, often using abstract rules, feedback, or response contingencies (Zühlsdorff et al., 2022; Dajani & Uddin, 2015; Diamond, 2013). Perseverative thinking measures, in contrast, capture how individuals experience their thoughts in daily life: whether thoughts feel repetitive, intrusive, uncontrollable, emotionally charged, or difficult to disengage from (Ehring & Watkins, 2008). The null findings for cognitive flexibility alongside the robust effects on perseverative thinking may therefore reflect a gap between “task-based flexibility” and “experienced flexibility.” Serotonergic interventions may not necessarily make individuals objectively better at switching between rules, but they may change how rigid or emotionally compelling negative thoughts feel. This interpretation is consistent with the broader literature on psychedelic microdosing and cognition. Pinhas et al. (2026) reported no significant overall effect of psychedelic microdosing on cognitive performance despite examining multiple cognitive domains and moderators. Importantly, that meta-analysis emphasized the value of focusing on behavioral cognitive tasks rather than self-report measures, precisely because subjective claims about cognitive enhancement can diverge from objective performance (Pinhas et al., 2026). The present findings extend this idea beyond microdosing: even when serotonergic full dose interventions are clinically or phenomenologically meaningful, their effects may not consistently appear in standard cognitive task performance.

The current findings are also in line with Doss et al. (2021) study of psilocybin therapy in major depressive disorder. Doss et al. reported improvements in cognitive and neural flexibility following psilocybin therapy, but these cognitive flexibility changes were not correlated with antidepressant improvement. This result is highly relevant to the present meta-analysis because it is suggested that even when cognitive flexibility improves after serotonergic psychedelic treatment, it may not be the central pathway through which symptoms improve. Together, these complementary findings raise the possibility that serotonergic interventions reduce depression and perseverative thinking through affective, experiential, or self-referential mechanisms rather than through a simple mediation of cognitive flexibility (Doss et al., 2021).

One alternative mechanistic pathway is that serotonin enhances mood and as a result enhances perseverative thinking. Perseverative thinking is strongly affective: rumination, worry, and obsessions are not merely cognitive habits, but emotionally loaded and often threat-related processes (Rachman, 1998; Nolen-Hoeksema et al., 2008; Newman et al., 2013). Serotonergic interventions may reduce negative affect, threat sensitivity, emotional reactivity, or the salience of intrusive thoughts (Harmer et al., 2017; Carhart-Harris & Nutt, 2017; Kraehenmann et al., 2015), and these changes may secondarily reduce perseverative thinking (Roiser et al., 2012). This possibility is particularly interesting because it suggests that perseverative thinking may not decrease because people become more cognitively flexible in a narrow executive-function sense, but because the emotional fuel that maintains repetitive thought is weakened. Future research should directly compare mediation models in which serotonin-related symptom-improvement is explained by changes in cognitive flexibility, mood, emotional reactivity, self-referential processing, or combinations of these pathways.

The distinction between serotonin depletion and serotonin elevation also deserves consideration. Acute tryptophan depletion reduces central serotonin availability by limiting precursor availability, whereas SSRIs increase serotonin availability primarily through serotonin transporter inhibition, and psychedelics act primarily through receptor-level mechanisms, especially 5-HT2A receptor agonism. These manipulations therefore are not mirror images of one another. A transient decrease in serotonin availability may not be sufficient to impair cognitive flexibility, particularly in healthy or mixed samples, while serotonin elevation may reduce perseverative thinking through mechanisms that are not captured by cognitive task performance. This distinction is important because serotonin does not function as a single mechanism. Different manipulations affect different parts of the serotonergic system and interactions with other systems such as dopamine, glutamate, and oxytocin (Daw et al., 2002; Vollenweider & Kometer, 2010; Mottolese et al., 2014).

The non-significant effect of serotonin-elevation on cognitive-flexibility may also reflect substantial methodological heterogeneity. Studies differed in task type (WCST, ID/ED, PRL, DRL, MAB etc.), dose (e.g., ranging widely from 5 to 300 mg/day across various SSRIs, from 1 mg microdoses up to 30 mg macro-doses for psilocybin, 50 to 100 µg for LSD, and 166 mg to 800 mg for daily tryptophan supplementation), timing of assessment (acute, chronic), and whether effects were measured within-subjects, between-subjects, pre-post, or relative to placebo. Cognitive flexibility is not a unform construct. Reversal learning, set-shifting, task-switching, creative flexibility, and cognitive control all depend on partly distinct neural and psychological processes (Dajani & Uddin, 2015). A serotonergic manipulation may affect one form of flexibility but not another or may even improve one aspect while disrupting another. Rather than showing a stable clustering of effects around zero, the estimates appear heterogeneous and directionally mixed. This suggests that the overall non-significant effect may result from averaging across studies that differ not only in effect size magnitude but also in direction. In this context, the null pooled estimate should not be interpreted as evidence that serotonin is unrelated to cognitive flexibility. Instead, it suggests that the available literature does not yet identify a consistent direction of effect across tasks, doses, designs, and serotonergic interventions.

These findings may be interpreted in light of Pinhas et al. (2026), who observed reduced cognitive control. The authors suggested that this pattern may align with theoretical accounts proposing that psychedelics relax top-down constraints on information processing. This interpretation is consistent with the REBUS model (Alamia et al., 2020; Carhart-Harris, 2018), which posits that psychedelics temporarily weaken high-level priors and rigid belief structures, thereby increasing cognitive flexibility. Importantly, the present findings showed no overall effect on cognitive performance while indicating reductions in rigid thinking patterns. Together, these results suggest that psychedelics may alter the style of cognition rather than its overall efficiency, reducing cognitive rigidity without necessarily improving or impairing objective cognitive performance.

On the neural level, the present findings can be interpreted through contemporary models of serotonergic modulation of large-scale brain networks. Classic psychedelics exert their effects primarily through agonism of cortical 5-HT2A receptors, which are densely expressed in regions involved in high-level integration and self-referential processing (Nichols, 2016). According to the REBUS model, psychedelics temporarily relax the precision of high-level priors, allowing rigid beliefs and entrenched patterns of interpretation to become more malleable (Alamia et al., 2020; Carhart-Harris, 2018). This mechanism may be particularly relevant to perseverative thinking, which is characterized by repetitive, inflexible cognitive patterns and excessive engagement of self-referential networks such as the default mode network (Hamilton et al., 2011). Within this framework, reductions in perseverative thinking may reflect a loosening of rigid self-related or threat-related beliefs rather than improvements in executive task performance. The present findings are consistent with this interpretation: serotonergic interventions were associated with reductions in rigid thinking patterns without corresponding improvements in objective cognitive performance. SSRIs may achieve similar reductions in perseverative thinking through a different pathway, gradually altering affective biases, emotional learning, and stress reactivity over time (Harmer et al., 2017). Thus, distinct serotonergic interventions may converge on reducing cognitive rigidity while operating through different temporal and neurobiological mechanisms.

Our results demonstrated that samples with a higher proportion of female participants showed larger reductions in perseverative thinking. This finding should be interpreted cautiously because sex was coded at the study/sample level rather than at the individual level. Nonetheless, it raises important questions. First, substantial evidence suggests sex differences in serotonergic functioning, including variation in serotonin synthesis, transporter availability, receptor expression, and hormonal modulation of serotonergic signaling (e.g., Jovanovic et al., 2008; Rubinow et al., 1998). In particular, fluctuations in estrogen and progesterone influence serotonin receptor sensitivity and transporter function, potentially altering responsiveness to serotonergic interventions. Second, women generally report higher baseline levels of rumination and other internalizing symptoms than men (Johnson & Whisman, 2013; Nolen-Hoeksema & Jackson, 2001). Because perseverative thinking is more prevalent in women, serotonergic interventions may have a larger therapeutic window for reducing these maladaptive cognitive patterns. Third, some evidence suggests sex differences in response to serotonergic treatments, with women showing greater responsiveness to SSRIs in certain contexts, potentially due to interactions between sex hormones and serotonin systems (Kornstein et al., 2000; Young et al., 2009). From the perspective of the present theoretical framework, these findings raise the possibility that women may be particularly sensitive to serotonergic modulation of cognitive rigidity and affective processing. Together, these results suggest that sex should not be treated merely as a demographic control variable, but as a theoretically meaningful moderator in serotonergic research.

We further observed different effects on different patterns of thinking. Perseverative thinking may not respond identically. Rumination did not significantly differ from obsessions, whereas worry showed a marginally smaller reduction relative to obsessions. This pattern should be interpreted cautiously, but it may suggest that serotonergic interventions are especially effective for forms of perseverative thinking that are more intrusive, repetitive, or compulsive, while worry may involve more future-oriented verbal problem-solving processes that are maintained by different mechanisms. More studies are needed to compare rumination, worry, and obsessions directly using harmonized measures.

An important strength of the present work is its integration of both objective cognitive outcomes and subjective symptom-related outcomes within a single serotonergic framework. Previous research has often examined either laboratory measures of cognitive flexibility or clinical symptoms in isolation, limiting the ability to evaluate whether improvements in subjective experience are accompanied by corresponding changes in cognition. By synthesizing evidence across both domains, the present study directly tests a central mechanistic assumption in affective neuroscience and psychopharmacology: that serotonergic interventions reduce maladaptive repetitive thought by enhancing cognitive flexibility. The findings challenge this assumption by demonstrating a dissociation between domains: serotonin elevation robustly reduced perseverative thinking while showing no consistent effect on objective cognitive flexibility tasks. This dissociation is theoretically informative because it suggests that laboratory-based measures of set-shifting and reversal learning may capture processes distinct from the cognitive-emotional flexibility relevant to daily life and psychopathology. Consequently, the study advances the field and provides a more nuanced account of serotonergic mechanisms, highlighting the importance of integrating behavioral and phenomenological outcomes when evaluating psychiatric interventions.

A further strength of the present work is its integration of multiple serotonergic manipulations within a single theoretical framework: acute tryptophan depletion (ATD), selective serotonin reuptake inhibitors (SSRIs), and classic psychedelics. These interventions are often studied in isolation because they target different components of the serotonergic system. Comparing these manipulations is scientifically valuable because they provide complementary windows into serotonergic function, allowing distinction between effects related to serotonin availability, receptor activation, and downstream neuroplastic adaptations. Such integration and juxtaposition of serotonin manipulation methods is particularly timely given the rapidly expanding role of serotonergic interventions in both research and clinical practice. SSRIs remain among the most widely prescribed psychiatric medications worldwide (Adjei et al., 2023), while psychedelic-assisted therapies are increasingly being investigated for depression, anxiety, obsessive-compulsive disorder, and other conditions characterized by cognitive rigidity and perseverative thinking (Chao & Horton, 2021; Collins, 2024). At the same time, experimental paradigms such as ATD continue to serve as critical tools for probing causal mechanisms of serotonin function (Young, 2013). By examining these approaches together, the present study moves beyond drug-specific effects and addresses a broader question of whether diverse serotonergic manipulations converge on common psychological processes. The finding that serotonin elevation consistently reduced perseverative thinking across both SSRIs and psychedelics suggests that different serotonergic interventions may converge on reducing cognitive rigidity through distinct neurobiological pathways. This integrative approach provides a more comprehensive understanding of serotonin’s role in psychopathology and is especially important in an era in which multiple serotonergic therapies coexist in both laboratory research and clinical care.

Several limitations should be considered when interpreting the present findings. The included serotonergic interventions differ substantially in their mechanisms of action. Acute tryptophan depletion, SSRIs, and psychedelics affect distinct components of the serotonergic system and therefore should not be interpreted as interchangeable manipulations of a single serotonergic mechanism. Although integrating these interventions provides a comprehensive perspective on serotonin, it also introduces conceptual heterogeneity. Similarly, cognitive flexibility is a multidimensional construct. The tasks included in this meta-analysis assessed partially distinct processes, including set-shifting, reversal learning, and decision-making under uncertainty. These paradigms rely on overlapping but non-identical cognitive and neural mechanisms, raising the possibility that serotonergic effects may be domain-specific and obscured when aggregated into a single meta-analytic estimate.

Moreover, substantial methodological heterogeneity was observed across studies, particularly in analyses of serotonin elevation. Studies varied in dosage, timing of assessment, study design, clinical population, and outcome measurement. Although multilevel models and robust variance estimation were employed, residual heterogeneity remained, suggesting the presence of unmeasured moderators. Additionally, most cognitive flexibility measures were laboratory-based tasks administered under highly structured conditions. Such measures may not adequately capture cognitive-emotional flexibility or the subjective experience of disengaging from repetitive negative thoughts in everyday life. The observed dissociation between objective and subjective outcomes may therefore partly reflect limitations of available measurement tools.

Notably, dose comparisons across serotonergic interventions should be interpreted cautiously. Unlike comparisons within a single drug class, there is no established method for converting or equating doses between SSRIs and psychedelics. Because these compounds differ substantially in their mechanisms of action, timing of administration, and downstream neurobiological effects, similar doses do not necessarily reflect comparable degrees of serotonergic modulation. Future research would benefit from the development of biomarkers or receptor occupancy measures that allow more direct comparisons across intervention classes.

In our third analysis of the effect of serotonin on perseverative thinking, we did not include acute versus chronic exposure as a separate moderator because exposure duration and specific serotonin manipulation were perfectly confounded in the available dataset. All psilocybin interventions were acute, whereas all SSRI interventions were chronic. This is not simply a statistical inconvenience; it reveals an important gap in the literature. Future studies should directly compare acute and repeated serotonergic interventions within the same pharmacological class, or examine the same intervention across multiple time windows.

In conclusion, this meta-analysis provides evidence for a dissociation between the effects of serotonergic manipulations on objective cognition and subjective perseverative thinking patterns. Neither acute serotonin depletion nor serotonin elevation produced consistent changes in laboratory measures of cognitive flexibility. In contrast, serotonin elevation was associated with robust reductions in pathological perseverative thinking, including rumination, worry, and obsessions across both SSRIs and classical psychedelics. These findings challenge prevailing assumptions that serotonergic interventions exert their therapeutic effects in part by enhancing executive cognitive flexibility. Instead, the present results support a more nuanced account in which serotonin may preferentially influence the phenomenological and affective dimensions of repetitive thought, reducing its emotional salience, self-relevance, and persistence, without necessarily altering performance on standard cognitive tasks. This distinction has important implications for theories of serotonin, suggesting that cognitive-emotional rigidity may constitute a more clinically relevant target than task-based flexibility. More broadly, these findings underscore the importance of integrating objective behavioral measures with subjective symptom outcomes when investigating neurobiological mechanisms of psychopathology and developing serotonergic treatments for disorders characterized by repetitive negative thinking.

## Data Availability

Data available upon request

